# Natural killer cell activation related to clinical outcome of COVID-19

**DOI:** 10.1101/2020.07.07.20148478

**Authors:** Christopher Maucourant, Iva Filipovic, Andrea Ponzetta, Soo Aleman, Martin Cornillet, Laura Hertwig, Benedikt Strunz, Antonio Lentini, Björn Reinius, Demi Brownlie, Angelica Cuapio Gomez, Eivind Heggernes Ask, Ryan M. Hull, Alvaro Haroun-Izquierdo, Marie Schaffer, Jonas Klingström, Elin Folkesson, Marcus Buggert, Johan K. Sandberg, Lars I. Eriksson, Olav Rooyackers, Hans-Gustaf Ljunggren, Karl-Johan Malmberg, Jakob Michaëlsson, Nicole Marquardt, Quirin Hammer, Kristoffer Strålin, Niklas K. Björkström, Karolinska COVID-19 Study Group

**Author notes:** Equal contribution (Christopher Maucourant, Iva Filipovic, Andrea Ponzetta).

## Abstract

Understanding innate immune responses in COVID-19 is important for deciphering mechanisms of host responses and interpreting disease pathogenesis. Natural killer (NK) cells are innate effector lymphocytes that respond to acute viral infections, but might also contribute to immune pathology. Here, using 28-color flow cytometry, we describe a state of strong NK cell activation across distinct subsets in peripheral blood of COVID-19 patients, a pattern mirrored in scRNA-seq signatures of lung NK cells. Unsupervised high-dimensional analysis identified distinct immunophenotypes that were linked to disease severity. Hallmarks of these immunophenotypes were high expression of perforin, NKG2C, and Ksp37, reflecting a high presence of adaptive NK cell expansions in circulation of patients with severe disease. Finally, arming of CD56bright NK cells was observed in course of COVID-19 disease states, driven by a defined protein-protein interaction network of inflammatory soluble factors. This provides a detailed map of the NK cell activation-landscape in COVID-19 disease.

## Introduction

The ongoing SARS-CoV-2 pandemic is presenting the human population with profound challenges. SARS-CoV-2 can cause COVID-19 disease, which in the worst cases leads to severe manifestations such as acute respiratory distress syndrome (ARDS), multi-organ failure, and death^1^. These manifestations may be caused by hyperactivated and misdirected immune responses. Indeed, high levels of IL-6 and an ensuing cytokine storm in the absence of appropriate type I and III interferon responses associate with severe COVID-19 disease^1-6^. Whereas emerging reports suggest that protective immunity is formed in convalescent patients with both neutralizing antibodies and SARS-CoV-2-specific T cells^7-9^, considerably less is known about early innate lymphocyte responses towards the SARS-CoV-2 infection and how they relate to host responses and disease progression. In this study, we evaluated natural killer (NK) cell activation in the context of SARS-CoV-2 infection and resulting COVID-19 disease.

NK cells are innate effector lymphocytes that are typically divided into cytokine-producing CD56bright NK cells and cytotoxic CD56dim NK cells^10^. Evidence for a direct role of NK cells in protection against viral infections comes from patients with selective NK cell deficiencies as these develop fulminant herpesviruses and other infections^11,12^. NK cells have also been shown to rapidly respond during the acute phase of several human infections including infections with hantavirus, tick-born encephalitis virus, influenza A virus (IAV), and dengue virus as well as during live-attenuated yellow-fever vaccination^13-16^. NK cells not only have the capacity to directly target and kill infected cells but can also influence adaptive T cell responses. Indeed, the degree of NK cell activation can function as a rheostat in regulating T cells where a certain level of NK cell activation might promote infection control while another degree of activation is related to immunopathology^17^. Compared to peripheral blood, the human lung is enriched for NK cells^16,18,19^. Human lung NK cells display a differentiated phenotype, and also have the capacity to respond to viral infections such as, e.g., IAV^16,18,19^. In COVID-19, emerging studies have reported low peripheral blood NK cell numbers in patients with moderate and severe disease^4,20-23^. Interestingly, two recent reports assessing the single cell landscape of immune cells in bronchoalveolar lavage (BAL) fluid of COVID-19 patients have suggested that NK cell numbers are increased at this site of infection^24,25^. However, a detailed map of the NK cell landscape in clinical SARS-CoV-2 infection has not been described yet.

Given the important role of NK cells in acute viral infections, their relatively high presence within lung tissue, as well as the links between NK cell activation, T cell responses, and development of immunopathology, we here performed a detailed assessment of NK cells in patients with moderate and severe COVID-19 disease. With strict inclusion and exclusion criteria, patients with moderate and severe COVID-19 disease were recruited early during their disease and 28-color flow cytometry-based analysis was performed focusing on assessment of NK cell activation, education, and expansion of adaptive NK cells. The results obtained are discussed in relation to the role of NK cell responses in acute SARS-CoV-2 infection and associated COVID-19 disease immunopathogenesis.

## Results

### Study design, clinical cohort, and general NK cell activation in COVID-19

Twenty-seven in-hospital patients with COVID-19 disease (10 moderate and 17 severe) were prospectively recruited after admission to the Karolinska University Hospital as part of a larger immune cell atlas effort (Karolinska COVID-19 Immune Atlas). Strict inclusion and exclusion criteria were used to ensure the setup of homogeneous patient cohorts, including time since symptom debut in relation to hospital admission, and disease severity staging (Fig. 1A, B). Seventeen healthy controls that were SARS-CoV-2 IgG seronegative and symptom-free at time of sampling were included as controls. To minimize inter-experimental variability and batch effects between patients and controls, all samples (n=44) were acquired, processed, and analyzed fresh on three consecutive weeks in April and May of 2020 at the peak of the COVID-19 pandemic in Stockholm, Sweden (Fig. 1B). See Material and Methods and Supplementary Table 1 and 2 for detailed patient characteristics.

**Figure 1.**
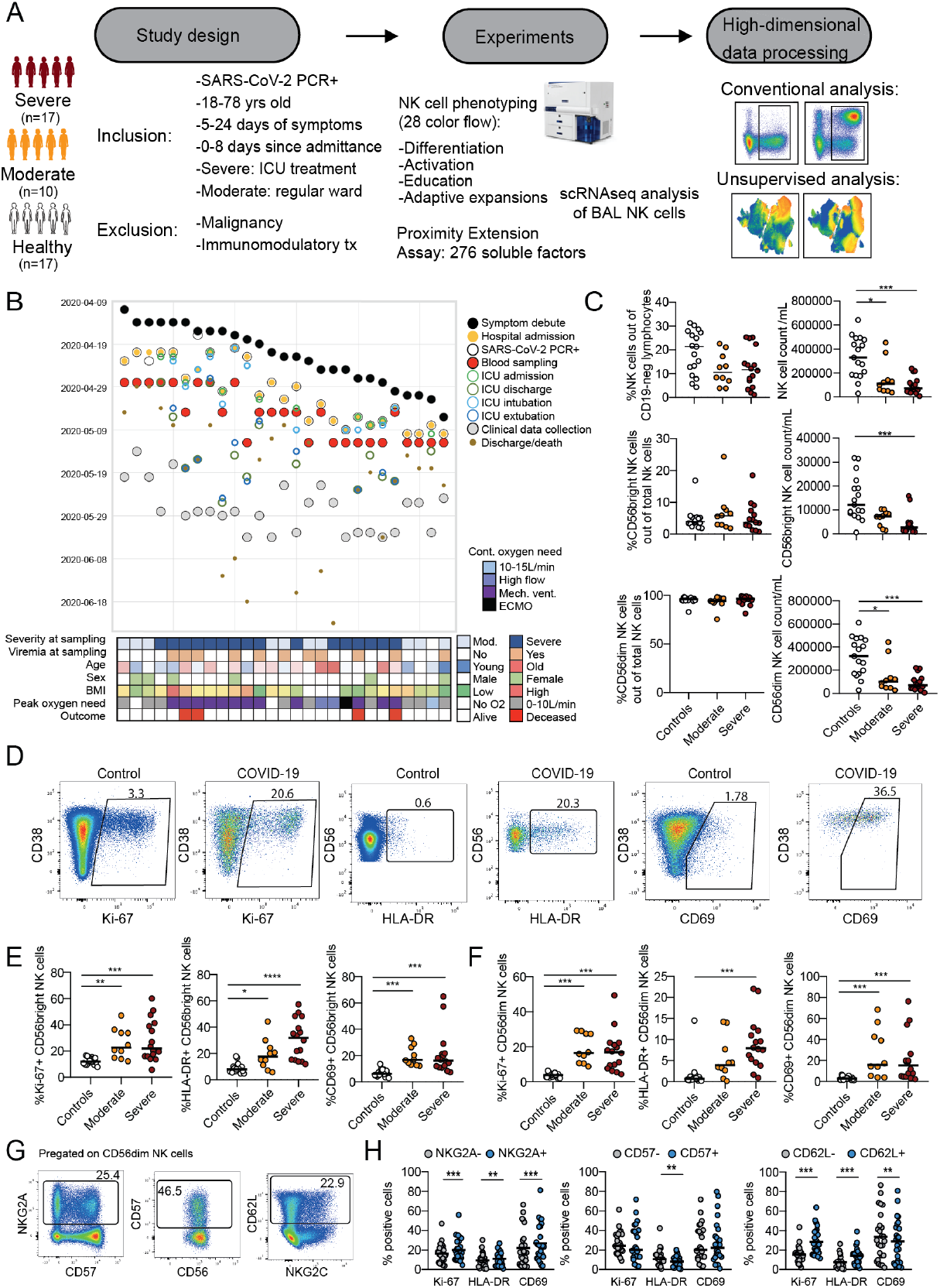
NK cells are robustly activated in moderate and severe COVID-19 disease. (A) Schematic overview of study design including main inclusion and exclusion criteria. (B) Swimmer plot illustrating the relationship between symptom debut, hospital admission, and blood sampling for the current study in relation to other main clinical events and clinical characteristics of included patients. (C) Summary data for percentage of NK cells and NK cell subsets as well as total NK cell and NK cell subset absolute counts for healthy controls (n=17), moderate (n=10), and severe (n=15-16) COVID-19 patients. (D) Representative flow cytometry plots for Ki-67, HLA-DR, and CD69 expression on NK cells in one healthy control and one COVID-19 patient. (e and f) Summary data for expression of the indicated markers in (E) CD56bright and (F) CD56dim NK cells in healthy controls, moderate and severe COVID-19 patients. (G) Representative flow cytometry plots or NKG2A, CD57, and CD62L expression on CD56dim NK cells. (H) Summary data for Ki-67, HLA-DR, and CD69 expression within NKG2A+/-, CD57+/, or CD62L+/- CD56dim NK cells from all moderate and severe COVID-19 patients (n=24-26). In C and E, statistical differences were tested using Kruskal-Wallis test followed by Dunn’s multiple comparisons test. In H, Wilcoxon matched-pairs signed rank test was performed. Numbers in flow cytometry plots indicate percentage of cells within a particular gate, bars represent median, *p < 0.05, ** p < 0.01, *** p < 0.001.

To study the NK cell response, peripheral blood mononuclear cells were analyzed with a 28-color NK cell-focused flow cytometry panel (Supplementary Table 3, gating strategy in Supplementary Fig 1). No change in total NK cell percentages, or the percentage of CD56dim and CD56bright NK cells was noted in patients compared to controls. However, the absolute count of total NK cells and the CD56dim and CD56bright subsets was severely reduced in patients compared to controls (Fig. 1C, Supplementary Fig. 1A). NK cell activation was assessed by analyzing expression of Ki-67, HLA-DR, and CD69 (Fig. 1D). A robust NK cell activation was noted in COVID-19 patients, where the degree of activation was similar in moderate and severe patients (Fig. 1E, F). CD56dim NK cells undergo continuous differentiation starting from less differentiated NKG2A+ and CD62L+ cells transitioning towards more terminally differentiated CD57+ NK cells^26,27^. A higher frequency of both NKG2A+ and CD62L+ CD56dim NK cells expressed Ki67 compared to NKG2A- and CD62L-cells in patients with COVID-19 (Fig. 1G, H). A subsequent Boolean analysis, simultaneously taking co-expression patterns of NKG2A, CD62L, and CD57 into account, revealed that the patients’ NK cell response occurred primarily among less differentiated CD62L-expressing NK cells (Supplementary Fig. 1B).

Taken together, using strict inclusion and exclusion criteria and patient recruitment during a defined period of time, we have obtained a well-controlled cohort of patients with moderate and severe COVID-19. Peripheral blood NK cells in these patients were robustly activated compared to healthy controls, but the general degree of activation was not associated with disease severity.

### Phenotypic assessment of NK cells in COVID-19

We next set out to perform a detailed phenotypic assessment of CD56bright and CD56dim NK cells in the COVID-19 patients. In a principal component analysis (PCA) of CD56bright (including 26 phenotypic parameters) and CD56dim NK cells (27 phenotypic parameters), patients clustered separately from healthy controls (Fig. 2A, B). This was, among other phenotypic markers, driven by changes in expression of CD98, Ki-67, and Ksp37 in CD56bright (Supplementary Fig. 2A), and Tim-3, CD98, CD38, CD69, and Ksp37 in CD56dim NK cells (Supplementary Fig. 2B). Conventional flow cytometry analysis further revealed increased expression of perforin, Ksp37, MIP-1*β*, CD98, Tim-3, and granzyme B in CD56bright NK cells from COVID-19 patients, as well as CD98, Tim-3, NKG2C, MIP-1*β*, and CD62L, among other markers, in CD56dim NK cells (Fig. 2C, D, Supplementary Fig. 2C, D). In line with the general activation profile (Fig. 1E, F), the PCA analysis did not highlight an obvious separation between moderate and severe COVID-19 patients (Fig. 2B) and similar results were obtained when unsupervised hierarchical clustering was performed on the expression of all studied phenotypic markers within CD56bright and CD56dim NK cells (Fig. 2E, F). Of note, the clustering clearly distinguished healthy from COVID-19 patients. Finally, the NK cell phenotype of responding (Ki-67 positive) compared to non-responding (Ki-67 negative) was compared. This analysis revealed that responding CD56bright NK cells co-expressed higher levels of granzyme B, CD25, HLA-DR, and Ksp37 as compared to non-responding cells and that responding CD56dim NK cells concurrently upregulated Tim-3 and HLA-DR (Supplementary Fig. 2E).

**Figure 2.**
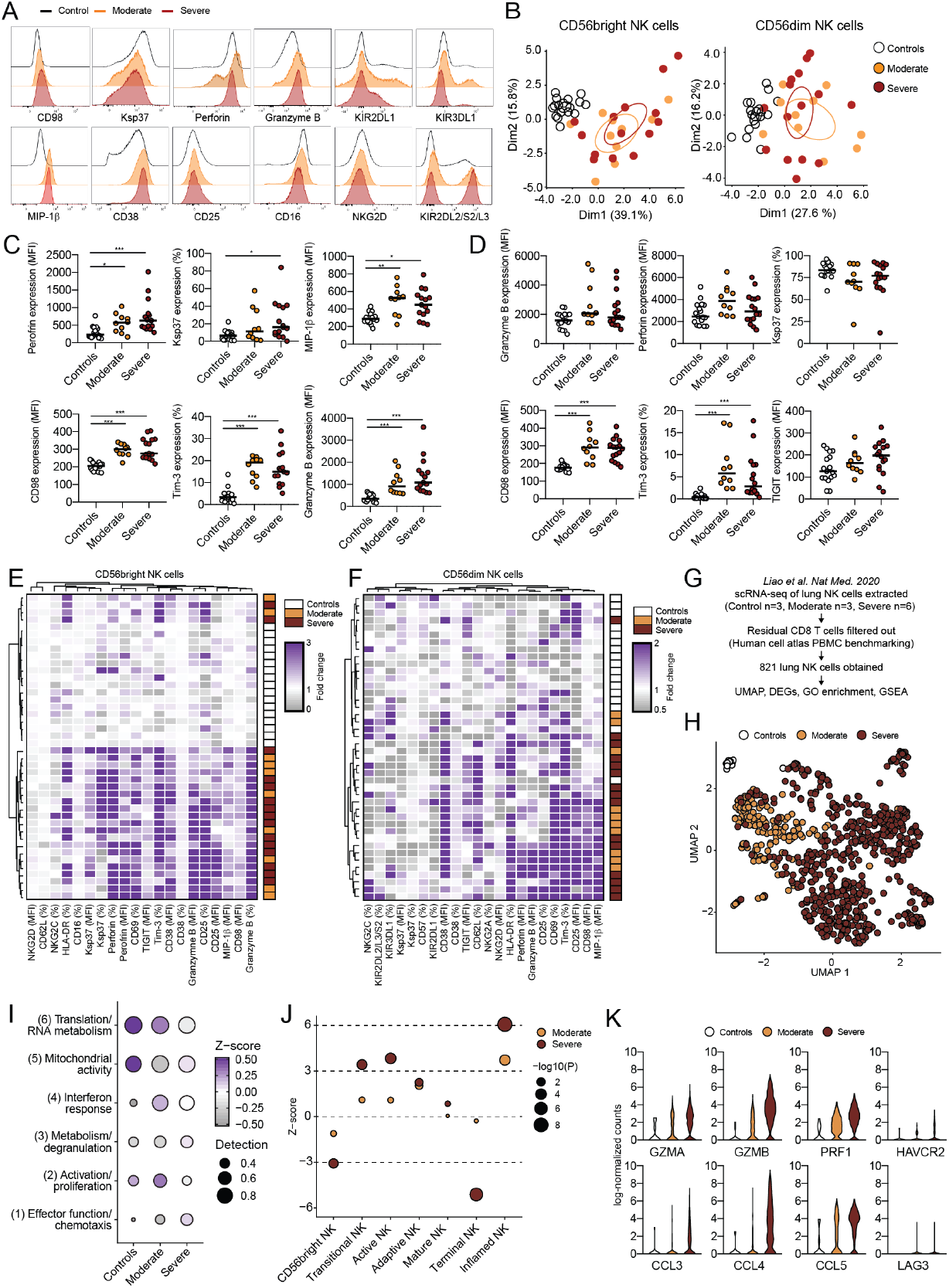
Detailed assessment of peripheral blood and lung NK cell activation in COVID-19 disease. (A) Representative flow cytometry histograms for the indicated markers on CD56dim NK cells in a healthy control, and moderate and severe COVID-19 patients. PCA of the phenotype of CD56bright and CD56dim NK cells in healthy controls, moderate, and severe COVID-19 patients. (C) Summary data for expression of the indicated markers on CD56bright NK cells in healthy controls (n=17), moderate (n=10), and severe (n=15) COVID-19 patients. (D) Summary data for expression of the indicated markers on CD56dim NK cells in healthy controls (n=17), moderate (n=10), and severe (n=16) COVID-19 patients. (E) Heatmap displaying fold change, after hierarchical clustering, in expression of the indicated markers (MFI or %) compared to median of the healthy control group for each studied individual (healthy control, moderate or severe COVID-19 patient) in (left) CD56bright and (right) CD56dim NK cells. Color indicate fold-change. (G) Strategy for analysis of single-cell RNA sequencing (scRNAseq) data obtained from BAL NK cells of controls and COVID-19 patients. (H) UMAP of scRNAseq data for BAL NK cells from controls, moderate, and severe COVID-19 patients. (I) Heatmap of indicated gene clusters obtained after gene ontology enrichment analysis on differentially expressed genes between the studied groups. Color indicate z-score, size of dots indicates detection level. (J) Z-score of indicated NK cell gene sets (obtained from scRNAseq of bone marrow NK cells) after gene set enrichment analysis. Size of dots indicate significance level using the Stouffer method compared to healthy controls. (K) Violin plots showing expression of the indicated genes, which were all differentially expressed from the pairwise comparison between controls, moderate, and severe COVID-19 patients. In C and D, statistical differences were tested using Kruskal-Wallis test followed by Dunn’s multiple comparisons test, bars represent median, *p < 0.05, ** p < 0.01, *** p < 0.001.

The activated status of NK cells in peripheral blood was also corroborated when analyzing publicly available single cell RNA sequencing data (scRNAseq) from NK cells in bronchoalveolar lavage fluid (BAL) from patients with COVID-19^24^ (Fig. 2G, H, Supplementary Fig. 2F). Clustering of differentially expressed genes (DEGs) in moderate and severe patients followed by gene ontology enrichment analysis of DEGs revealed six distinct gene modules where effector functions, activation/proliferation, and interferon response were enriched in patients compared to controls (Fig. 2I, Supplementary Fig. 2G, H, Supplementary Table 4). Intriguingly, the interferon response and activation/proliferation signatures were most highly enriched in BAL NK cells from patients with moderate disease whereas severe patients had a higher effector function gene signature (Fig. 2I, Supplementary Fig. 2G, H). Gene set enrichment analysis further highlighted that NK cells from BAL fluid of severe patients displayed an activated and inflamed profile (Fig. 2J, Supplementary Fig. 2I). Finally, several of the proteins that were increased in peripheral blood NK cells from COVID-19 patients (Fig. 2C-F, Supplementary Fig. 2C, D) were also identified as DEGs in lung NK cells of COVID-19 patients as compared to controls, including *GZMB* (granzyme B), *PRF1* (perforin), *HAVCR2* (Tim-3), and *CCL4* (MIP-1*β*) (Fig. 2K).

In summary, peripheral blood CD56bright and CD56dim NK cells displayed an activated effector phenotype that is similar in nature in moderate and severe COVID-19 disease, and that could be corroborated in analysis of NK cells from BAL fluid of COVID-19 patients.

### Impact of KIR expression and NK cell education on the NK cell COVID-19 response

CD56dim NK cell function is regulated by inhibitory receptors where subsets of these cells expressing inhibitory receptors with cognate self MHC class I ligands present in the host are educated and thus, more functional^28,29^. We next evaluated the impact of expression of inhibitory receptors, KIRs and NKG2A, as well as of NK cell education on the acute COVID-19 response (Supplementary Fig. 3A, Supplementary Table 5). No significant differences were found in the fractions of NK cells expressing different combinations of inhibitory KIRs and NKG2A in COVID-19 patients as compared to controls (Supplementary Fig. 3B). Furthermore, the size of CD56dim NK cell subsets being educated via KIRs or NKG2A was also similar in patients and controls (Supplementary Fig. 3C). Finally, we assessed the NK cell response by measuring Ki-67 upregulation in CD56dim NK cell subsets, either solely based on KIR and NKG2A expression or also by integrating information on MHC class I ligand haplotypes (Supplementary Fig. 3D-F). This comparison indicated that the NK cell response in acute COVID-19 occurred independently of inhibitory KIR expression and NK cell education status.

### Adaptive NK cell expansions in severe COVID-19

Our analyses so far revealed robust activation of NK cells with an effector phenotype in COVID-19 patients. The response appeared independent of disease severity as few differences were observed when comparing moderate and severe patients. However, we noticed that severe patients had increased frequencies of NKG2C+ NK cells (Supplementary Fig. 2D). This indicated the presence of adaptive NK cell expansions, which are often associated with CMV infection and can be found activated when CMV-seropositive patients undergo other severe acute viral infections^13,30,31^. Thus, we next evaluated the presence of adaptive NK cell expansions in COVID-19 patients by gating on NKG2C+CD57+ NK cells and assessed their expression of KIRs, NKG2A, and CD38 (Fig. 3A, B, Supplementary Fig. 4A, B). A higher percentage of CD56dim NK cells from severe COVID-19 patients co-expressed NKG2C and CD57 (Fig. 3C), and while the absolute number of NKG2C-CD57-cells decreased in COVID-19 patients, NKG2C+CD57+ CD56dim NK cells remained stable even in patients with severe disease course (Fig. 3D, E). Adaptive NK cell expansions were confined to CMV seropositive individuals (Fig. 3F and Supplementary Fig. 4A). Our strategy to identify expansions in the smaller healthy control cohort showed good correspondence with a previous study assessing these in a large cohort of more than 150 healthy individuals (Fig. 3G). Strikingly, approximately two-thirds of CMV seropositive severe COVID-19 patients had such an expanded adaptive NK cell population compared to one third in controls and even fewer in the moderate COVID-19 patients (Fig. 3F-H). Notably, the clinical status was similar in severe patients with and without adaptive NK cell expansions (Supplementary Fig. 4F-H, Supplementary Table 6A, B).

**Figure 3.**
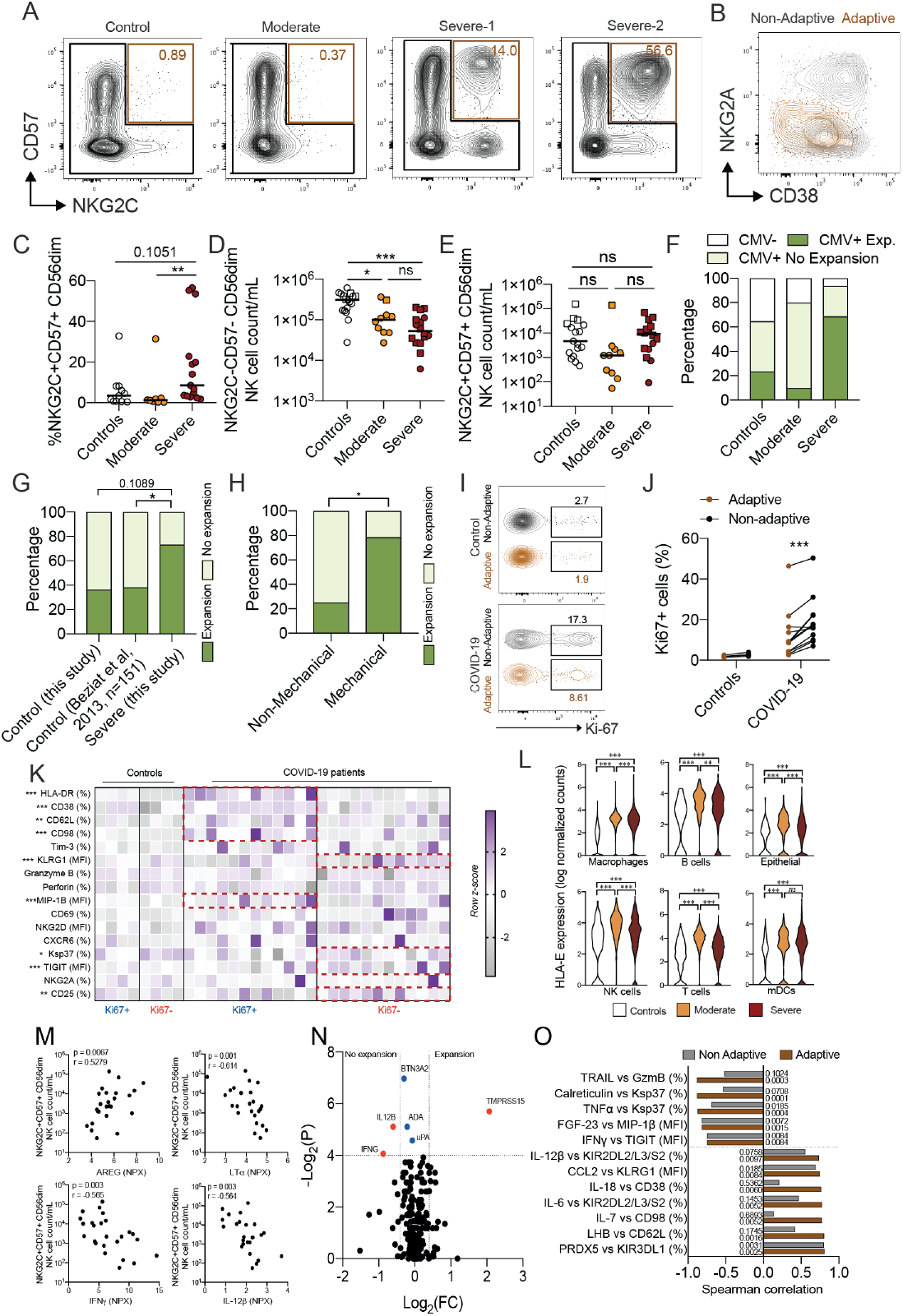
Adaptive NK cell expansions associate with severe COVID-19 disease. (A) Representative flow cytometry plots for CD57 and NKG2C expression on CD56dim NK cells in one healthy control, one moderate, and two severe COVID-19 patients. (B) Representative flow cytometry plot for NKG2A and CD38 expression for adaptive (NKG2C+CD57+, brown) and non-adaptive (non-NKG2C+CD57+, black) CD56dim NK cells. (C) Summary data for frequency of the indicated subsets in CMV seropositive healthy controls (n=11), moderate (n=8), and severe (n=15) COVID-19 patients. (D-E) Absolute counts of the indicated subsets in healthy controls (n=17), moderate (n=10), and severe (n=16) COVID-19 patients. Squares represent individuals with NK cell adaptive expansions while circles represent those without expansions. (F-H) Percentage of individuals from the indicated groups having or not having adaptive NK cell expansions. (I and J) Representative flow cytometry plots and summary data for Ki-67 expression in adaptive and non-adaptive CD56dim NK cells in healthy controls (n=4) and COVID-19 patients (n=12). (K) Heatmap displaying the expression z-score (see Methods) of indicated markers (MFI or %) in proliferating/resting adaptive NK cells from healthy controls (n=4) and COVID-19 patients (n=12). Rows were ranked from top to bottom by decreasing z-score in Ki67+ adaptive NK cells in COVID-19 patients. Red boxes highlight the markers with increased expression in either Ki67+ or Ki67-adaptive NK cells. (L) Violin plots showing gene expression of *HLA-E* from scRNAseq data of the indicated lung immune and non-immune cell types in healthy controls, moderate, and severe COVID-19 patients. FDR from pairwise comparisons (see Methods) are shown for statistical significance. (M) Spearman correlations between the absolute count of NKG2C+CD57+ CD56dim NK cells and the indicated soluble factors in moderate and severe COVID-19 patients. (N) Volcano plot showing soluble factors that are specifically increased or decreased in severe COVID-19 patients with or without adaptive NK cell expansions. (O) Spearman correlations, with relative p-values, between the expression of indicated markers on adaptive (brown) or non-adaptive (grey) NK cells from matched COVID-19 patients and the indicated soluble factors. In C-E, statistical differences were tested using Kruskal-Wallis test followed by Dunn’s multiple comparisons test, in F-H using Fisher’s exact test and in j-k using Wilcoxon matched-pairs signed rank test. Bars represent median, *p < 0.05, ** p < 0.01, *** p < 0.001.

The adaptive NK cell expansions found in severe COVID-19 patients displayed signs of proliferation, although at lower levels compared to non-adaptive NK cells (Fig. 3I, J, Supplementary Fig. 4C-E). Responding adaptive NK cells expressed HLA-DR, CD38, CD62L, and MIP-1*β* at higher frequencies, but lower levels of Ksp37, TIGIT, and CD25 as compared to non-responding adaptive NK cells (Fig 3K, Supplementary Fig. 4I). Next, we set out to more specifically address what was driving the emergence of adaptive NK cell expansions. Re-analyses of published scRNAseq data revealed that both immune and non-immune cell types displayed upregulated expression of genes encoding *HLA-E* and *HLA-A, B*, and *C* expression in BAL fluid of moderate and severe COVID-19 patients as compared to controls (Fig. 3L and Supplementary Fig. 4J). The number of NKG2C+CD57+ NK cells was inversely correlated with several soluble factors, including LT*α*, IL-12*β*, IFN*γ*, and amphiregulin (AREG) (Fig. 3M). Similarly, when specifically comparing soluble factors in severe patients with and without expansions, only a limited number of factors were differentially expressed (Fig. 3N). In line with this, associations between phenotypic features and soluble factors were in most cases similar for adaptive and non-adaptive NK cells (Fig. 3O).

Taken together, severe, but not moderate, COVID-19 disease is associated with a high presence of adaptive NK cell expansions that display signs of proliferation and activation.

### Dimensionality reduction analysis of NK cells separates severe from moderate patients

Except for adaptive NK cell expansions being more prevalent in severe COVID-19 patients, manual gating-based flow cytometry analysis did not identify features specific to either moderate or severe COVID-19 patients. In an attempt to identify these features through an unsupervised approach, we performed Uniform Manifold Approximation and Projection (UMAP) analysis on all patients and controls (Fig. 4A, B, Supplementary Fig. 5). This revealed distinct topological regions between patients and controls (Fig. 4A). We applied PhenoGraph to our samples to further describe NK cell subpopulations and to quantify differences in their relative abundance between COVID-19 patient groups. PhenoGraph clustering rendered 36 distinct clusters (Fig. 4C). Approximately half of these clusters were equally shared between controls, and moderate and severe patients (Fig. 4D), and some clusters were patient specific. Furthermore, this analysis also revealed clusters that contained NK cells predominately from controls, moderate, or severe COVID-19 patients (Fig. 4C, D). The phenotype of NK cells in clusters with similar relative abundance across the three groups was homogenous, but greater differences were observed when assessing clusters uniquely and significantly enriched in moderate or severe patients (Fig. 4E, F, Supplementary Fig. 5D).

**Figure 4.**
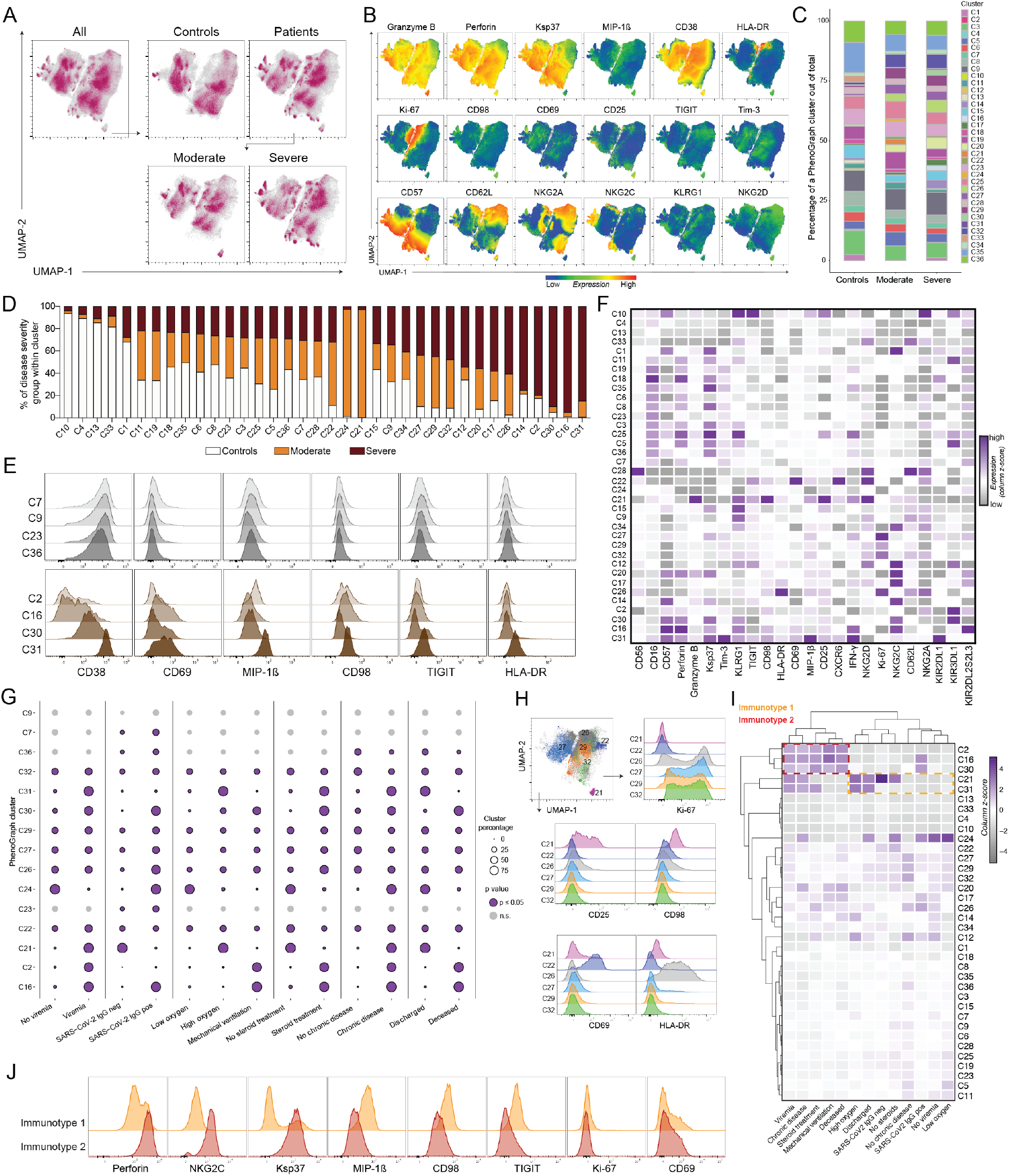
Automated analysis of NK cells in COVID-19 identifies putative NK ‘immunotypes’ differentially enriched in two main groups of patients. (A) UMAP of all events, of controls and patients, and then of patients split into moderate and severe group. (B) Representative expression of phenotypic markers on all cells in the UMAP. (C) The percentage of all identified 36 PhenoGraph clusters within total cells of controls, moderate, and severe COVID-19 patients. (D) Relative abundance of control, moderate, and severe groups within each PhenoGraph cluster. (E) Expression of selected markers from representative PhenoGraph clusters that did not display significant differential relative abundance between healthy, moderate, and severe groups (top histograms) and four significant clusters that were most highly abundant in severe COVID-19 patients (bottom histograms). (F) Heatmap showing expression of all phenotypic markers in all identified PhenoGraph clusters. Data shown represents column z-score of median expression values. (G) Balloon plot showing the percentage of NK cell clusters from COVID-19 patients stratified based on the indicated clinical categorical parameters. Purple circles with a border indicate significant PhenoGraph clusters in a particular comparison (p ≤ 0.05) and light-grey circles without a border indicate non-significant clusters (Materials and Methods, Supplementary Table 7). (H) Selected PhenoGraph clusters overlaid on the UMAP as well as representative flow cytometry histograms showing staining of the indicated markers within the clusters. (I) Hierarchical clustering of PhenoGraph clusters and clinical categorical parameters. Heatmap was calculated as a column z-score of cluster percentages. Putative NK immunotypes are indicated (Immunotype 1 and Immunotype 2). (J) Histograms showing expression of separating and non-separating markers within the NK cell immunotype 1 and 2 clusters.

Next, each PhenoGraph cluster was stratified within patients based on six clinical categorical parameters (viremia, seroconversion, oxygen need, steroid treatment, underlying comorbidities, and outcome) (Fig. 4G) in the same manner as moderate or severe patient categories were defined (Materials and Methods). This revealed that most clusters showed a significantly different relative abundance between patient categories and healthy controls in at least one of the clinical parameter-defined patient groups (Fig. 4G, Supplementary Fig. 6A, Supplementary Table 7). Importantly, for some clusters, the relative abundances were different depending on the clinical parameter. This approach was critical for our understanding, as it enabled us to step away from looking at disease severity on the whole and to instead look into the defining clinical parameters separately. Cluster 21, with high CD25 and CD98 expression (Fig. 4H), was almost completely absent from patients who were on mechanical ventilation and the ones who died from COVID-19, for example. Conversely, CD69-expressing C22, and the four most highly Ki-67-expressing clusters accounted for a similar percentage across all patient categories in our dataset (Fig. 4H, Supplementary Fig. 6B-F). Hierarchical clustering of PhenoGraph clusters’ relative abundance across clinical categorical parameters revealed the presence of two “immunotypes” in patients, one enriched categorical parameters attributed to milder disease course whereas the other one contained for viremia, chronic underlying diseases, steroid treatment, mechanical ventilation, and fatal outcome (Fig. 4I). Indeed, a similar clustering was obtained when moderate and severe disease status was included (Supplementary Fig. 6G). Finally, the NK cell phenotypes associated with moderate (immunotype 1) versus severe (immunotype 2) disease were assessed. This revealed upregulated MIP-1*β*, CD98, and TIGIT in the moderate immunotype and conversely high expression of perforin, NKG2C, and Ksp37 in the severe immunotype whereas the activation markers Ki67 and CD69 remained unchanged (Fig. 4J). Interestingly, high perforin, NKG2C, and Ksp37 are features of adaptive NK cell expansions which were also associated with severe disease (Fig. 3).

In summary, unsupervised high-dimensional analysis of the NK cell response in COVID-19 disentangles NK cell phenotypes associated with disease severity.

### CD56bright NK cell activation associate with disease severity

Finally, we performed a more detailed analysis of the NK cell response in relation to clinical laboratory parameters and clinical scoring systems. As expected, a PCA of moderate and severe patients based on clinical laboratory parameters revealed separation between the two groups (Fig. 5A). This was primarily driven by high systemic levels of neutrophils, IL-6, D-dimer, and c-reactive protein, all known clinical laboratory data typically associated with severe disease (Fig. 5B). Out of the altered NK cell phenotypic parameters (Fig. 2, Supplementary Fig. 2), the expression levels of perforin and granzyme B on CD56bright NK cells correlated with IL-6 levels in the patients (Fig. 5C). Indeed, perforin on CD56bright NK cells in the COVID-19 patients also positively correlated with Sequential Organ Failure Assessment (SOFA) and SOFA-R scores as well as neutrophil count and negatively with PaO2/FiO2 ratio (Fig. 5D), all in line with this NK cell phenotype associating with disease severity. The upregulation of perforin and granzyme B on CD56bright NK cells (Figure 2) was further positively associated with a general activation and upregulation of effector molecules within CD56bright and CD56dim NK cells and inversely correlated with expression of the inhibitory check-point molecule TIGIT (Fig. 5E, Supplementary Fig. 7A and B). As such, the association with high perforin was in line with the severe immunotype identified through unsupervised high-dimensional analysis (Fig. 4).

**Figure 5.**
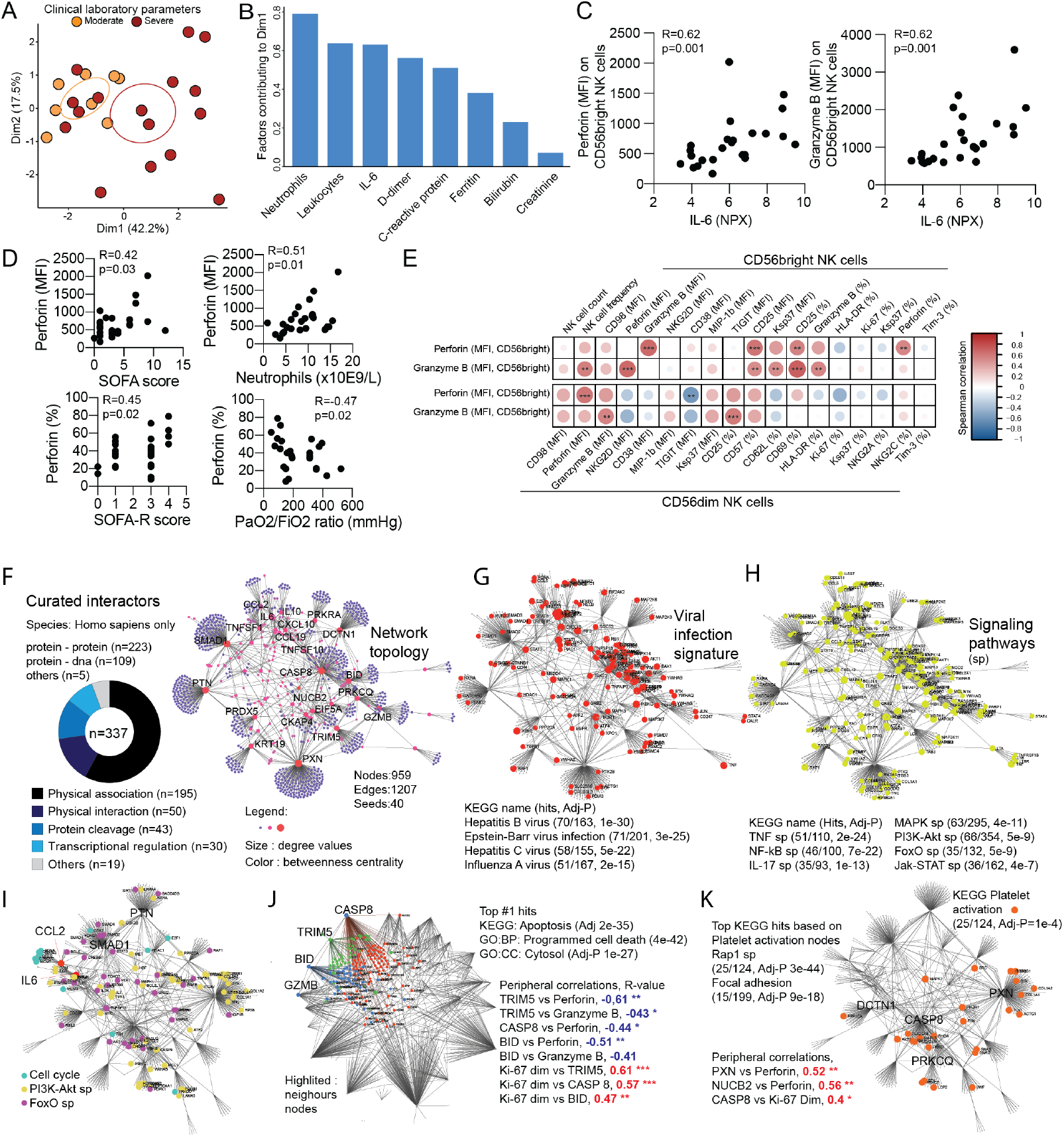
Arming of CD56bright NK cells associate with COVID-19 disease severity. (A) PCA of COVID-19 patients based on clinical laboratory parameters. (B) Bar plot showing the clinical laboratory parameters that contributed to dimension 1 (dim 1) in Figure 5A. (C) Spearman correlations between the indicated CD56bright NK cell phenotypic parameters in COVID-19 patients and serum IL-6 levels (n=24). (D) Spearman correlations between the indicated CD56bright NK cell phenotypic parameters and clinical parameters (n=25). (E) Correlation matrix showing spearman correlations between perforin and granzyme expression (MFI) on CD56bright NK cells and the indicated other NK cell phenotypic parameters (n=25). Color indicate R-value, and asterisks indicate p-values. (F) Topology and content of the protein-protein interaction network driven by soluble factors (seeds) correlated with perforin and granzyme B expression in CD56bright NK cells. Superimposition of nodes involved in (G) viral infection and (H) signaling pathways on the identified protein-protein interaction network. (I) FoxO, PI3K-Akt, and cell cycle pathways. (J) Neighbor nodes of Caspase-8, TRIM5, BID, and associated KEGG pathways. (K) Platelet activation and associated KEGG pathways.

Thus, integration of clinical laboratory parameters into phenotypical NK cell analysis revealed that arming of CD56bright cells with cytotoxic molecules correlated with parameters associated to a severe disease course.

### Protein-protein interaction network driving CD56bright NK cell activation in COVID-19

To better understand the significance of the activated CD56bright NK cell phenotype and its association with disease severity we took advantage of an extensive characterization of soluble serum proteins that had been performed on the same 27 patients through the Karolinska COVID-19 Immune Atlas effort. Of the 276 quantified soluble factors, the concentrations of 58 were significantly associated with perforin and granzyme B expression on CD56bright cells and 9 showed strong, primarily positive, correlations (Supplementary Fig. 7C). To gain insight into the biological basis of the peripheral protein signature that correlated with CD56bright NK cell activation, we analyzed protein-protein interaction networks. We identified 337 interactions (Supplementary Table 8A) that were integrated into a network of 959 elements (nodes, Fig. 5F, Supplementary Table 8B). Many of these related to viral infections, suggesting redundancy between different viral infections in NK cell activation mode (Fig. 5G, Supplementary Table 8C). Additionally, several distinct signaling pathways potentially involved in the CD56bright NK cell response in COVID-19 were identified (Fig. 5H, Supplementary Table 8C). The contribution of these pathways to cell proliferation was further dissected, identifying IL-6 as part of the Pi3K-Akt and FoxO pathways as well as CCL2 (Fig. 5I). Interestingly, TRIM5, Caspase-8, and BID were all positively correlated with NK cell proliferation but negatively with granzyme B expression (Fig. 5J). These three soluble factors were part of the same biological process of apoptosis and cell death (Fig. 5J). Finally, we identified a significant representation of proteins associated with platelet activation (Fig. 5K). These proteins also pointed towards a role for Rap1 signaling (integrin-mediated cell adhesion) and focal adhesion (involving cellular cytoskeleton and extracellular matrix) in the identified network between CD56bright NK cell arming in COVID-19 and soluble factors.

Altogether, our findings indicate that NK activation pathways could be shared with other viral infections, connecting NK cell-specific phenotypic traits to components of the systemic inflammatory milieu related to COVID-19 disease pathogenesis.

## Discussion

Using high-dimensional flow cytometry, integration of soluble factors analysis, and complementing with analysis of scRNAseq data, we here characterized the host NK cell response towards SARS-CoV-2 infection in moderate and severe COVID-19 patients in the systemic circulation. Results revealed low NK cell numbers but a robustly activated NK cell phenotype in COVID-19 patients. Peripheral blood NK cell activation states were mirrored in transcriptional signatures of BAL NK cells of COVID-19 patients from another cohort. Unsupervised high-dimensional analysis furthermore revealed clusters of NK cells that were linked to disease status. Finally, severe hyperinflammation, defined by clinical parameters, was associated with a high presence of adaptive NK cell expansions and arming of CD56bright NK cells. These systemic immune changes were linked to a soluble factor protein interaction network displaying a canonical viral activation signature as well as distinct signaling pathways. Altogether, the results provide a comprehensive map of the NK cell response landscape in SARS-CoV-2 infected COVID-19 patients and yield insights into host response reactions towards viral infections and associated disease pathology.

NK cells are known to rapidly respond during diverse acute viral infections in humans including those by dengue virus, hantavirus, tick-borne encephalitis virus, and yellow fever virus^13-15,32^. Although a similarly detailed analysis of NK cells has not been performed in acute SARS-CoV-2 infection causing COVID-19, early reports from the pandemic have indicated low circulating NK cell numbers in patients with moderate and severe disease^20,21,23^. Those reports are in line with what is reported here. Transiently reduced NK cell numbers during the acute phase of infections have also been shown in SARS-CoV-1 infection^33^ and in acute hantavirus infection^13^. The magnitude of the present detected NK cell response, with a quarter of the circulating cells either displaying signs of proliferation or activation, mirrors the responses observed during acute dengue fever as well as in acute hantavirus infection^13,14^. The present in-depth phenotypic assessment of NK cells in moderate and severe COVID-19 patients revealed an activated phenotype with upregulated levels of effector molecules and chemokines, activating receptors, and nutrient receptors. We also observed signs of inhibitory immune check-point receptor upregulation through increased levels of TIGIT and Tim-3. As such, the present results confirms and extends, at protein level, what was recently reported for peripheral blood NK cells using single-cell RNA sequencing^20^.

Human NK cells are enriched in lung compared to peripheral blood^18,19^. COVID-19 patients display significant immune activation in lungs, where moderate disease is associated with a high number of T cells and NK cells while severe disease patients’ lungs, on the other hand, are enriched in neutrophils and inflammatory monocytes^24,25,34^. By analyzing publicly available scRNAseq data on BAL NK cells from COVID-19 patients^24^, these cells were found to be strongly activated. Interestingly, the interferon response appeared strongest in BAL NK cells from moderate COVID-19 patients. This is in line with severe COVID-19 associating with a blunted interferon response^6^. We further confirmed that a similarly activated profile of NK cells was present in BAL fluid as was in systemic circulation, including high expression of effector molecules and chemokines. Altogether, this suggests that NK cells have a role in the early host immune response towards SARS-CoV-2 at the site of infection.

Under homeostatic conditions, NK cell functional responses are gradually acquired during differentiation^26^ and upon engagement of NKG2A and other inhibitory receptors recognizing self-MHC-I molecules, in a process known as education^28,29^. In the present cohort, we did not detect differences in KIR expression, including those binding self-MHC-I molecules, in moderate or severe COVID-19 patients. This finding is consistent with a previous report on dengue virus infection where the global NK cell receptor repertoire was not altered upon infection^14^, and significantly diverges from what has been reported for mouse CMV, where the uneducated fraction represents by far the most responsive NK cell subset^35^. Moreover, while active human CMV infection is known to significantly impact the global NK cell KIR-receptor repertoire through the expansion of NKG2C+ cells that preferentially express educating KIRs^36^, the present results indicate that this may not occur during SARS-CoV-2 infection, at least in peripheral blood. Together, the findings suggest that the strong response shown by circulating NK cells identified in COVID-19 patients does not depend on education.

This is the first time an increase of adaptive NK cell expansions is described in COVID-19 patients. Strikingly, this feature was almost exclusively found in SARS-CoV-2 infected patients with severe disease. The frequency of adaptive NK cell expansions in the present healthy control cohort was comparable with previous findings on larger healthy CMV-seropositive human cohorts^36,37^, strengthening our observations in COVID-19 patients. While a recent study reported an increase in NKG2C expression and a corresponding decreased expression of the adaptive NK cell-related marker FcεR1γ^38^, we here provide a deep dissection of the adaptive NK cell phenotype, highlighting that despite their lower responsiveness to cytokines^37^, a significant percentage of adaptive NK cells (up to 45%) actively proliferate in COVID-19 patients, albeit at a lower rate compared to non-adaptive NK cells. Moreover, in stark contrast with the robust numeric reduction observed of non-adaptive NK cells as well as in many other lymphocyte subsets during acute SARS-CoV-2 infection^39,40^, adaptive NK cells were quantitatively unchanged (or even increased) in severe COVID-19 patients. This suggests that their relative accumulation in blood in course of SARS-CoV-2 infection might be related to their higher resistance to cytokine-induced apoptosis^37^ or to a different sensitivity to chemotactic signals compared to non-adaptive NK cells.

Given their high capacity to perform antibody-dependent cell cytotoxicity (ADCC)^37^, it will be important to determine whether adaptive NK cells could play a protective role against SARS-CoV-2 infection upon production of virus-specific antibodies reported in convalescent COVID-19 patients^41-43^. On the other hand, a quantitative or qualitative defect in neutralizing antibody production might lead to increased viral pathogenesis and to detrimental effects, in a process known as antibody-dependent enhancement^44^. Except for ADCC, adaptive NK cells can also sense target cells via NKG2C directly recognizing HLA-E. Here, we found upregulated *HLA-E* in immune and stromal cells in BAL fluid of COVID-19 patients. In contrast, the expression levels of both *HLA-E* as well as genes encoding other MHC class I molecules were recently reported to be downregulated in peripheral blood immune cells from COVID-19 patients^20^, suggesting that a receptor-ligand driven expansion of adaptive NK cells might occur *in situ* rather than in circulation. Expansion of adaptive NK cells in CMV-seropositive patients was previously reported in acute hantavirus infection^13^, and was associated with the upregulation of both class I MHC and HLA-E on virus-infected cells. The absence of strong links between soluble factors and adaptive NK cell expansions suggest that their phenotype is instead driven by receptor-ligand interactions in COVID-19. Indeed, in CMV infection, virus-encoded peptides presented on HLA-E serve as a key activator of NKG2C+ cells and, together with cytokines, contribute to their expansion and differentiation^45^. Interestingly, the potential relevance of the NKG2C-HLA-E axis in the context of SARS-CoV-2 infection is highlighted by a recent study showing an increased prevalence of the allelic variant NKG2C^del^, associated with a reduced NKG2C expression, among severe COVID-19 patients, compared to the healthy population^46^. Moreover, a higher frequency of the HLA-E*0103 allele was found in ICU-hospitalized patients from the same cohort. Taken together, our working hypothesis is that a higher frequency of NKG2C+ NK cells could be functionally required in order to unleash NK cell antiviral activity in SARS-CoV-2 infection, particularly in more severe patients, while genetic predisposition leading to the reduction of NKG2C could cause more severe clinical manifestations.

We included viremia and other clinical categorical data that associate with disease severity in the UMAP analysis of NK cells in COVID-19 patients. These integrated immune signatures allowed us to, without *a priori* knowledge of the disease stage, identify two immunotypes associated with COVID-19 severity. The immunotype that was linked to severe disease displayed high expression of perforin, Ksp37, and NKG2C. This corroborated our finding of increased adaptive NK cells in severe COVID-19 since a link between NKG2C and disease severity was revealed in two independent types of analyses. Similarly, both perforin and Ksp37 were part of the compound phenotype of armed CD56bright NK cells that we found associated with severe disease and the scRNAseq analysis identified effector function as the gene ontology module most highly detected in severe patients. The immunotype linked to moderate disease, on the other hand, was associated with higher expression of MIP-1*β*, CD98, and TIGIT. The association between high expression of the inhibitory checkpoint TIGIT and moderate disease was strengthened by the fact that the armed CD56bright NK cell phenotype negatively correlated with TIGIT expression. Future work should address if containment of NK cells by inhibitory checkpoints could aid in containing hyperinflammation and severe COVID-19 disease.

A strong association between arming of NK cells and disease severity was observed, which was coupled to IL-6 levels but also to several clinical disease severity scores. Our protein-protein network linked to this arming revealed that interactions active in COVID-19 were shared with other viral infections such as hepatitis B and influenza A virus. Furthermore, IL-6 associated with Pi3K-Akt and FoxO signaling in NK cells. Both of these pathways are known to coordinate the cell cycle and have previously been reported to be activated in NK cells during acute dengue infection^14^. The analysis also revealed a role for CCL2 in cell cycle regulation, possibly reflecting a shared role for this factor in activation and migration of both NK cells^47-49^ and myeloid cells^24,25^ to the site of infection^50^. Apoptosis and cell death pathways, with TRIM5, Caspase-8, and BID, were further identified and linked in a reciprocal manner to NK cell arming (negative) and proliferation (positive). BID and Caspase-8 are effector molecules of the apoptotic pathway triggered by Fas or TRAIL^51^ whereas TRIM5 is an ISG targeting post-entry stages of retrovirus replication^52^. This could suggest that a balance exists between NK cell activation and proliferation on one hand and direct effector function on the other hand. Finally, the network analysis also revealed an overrepresentation of interactions relating to platelet activation, which was reported to contribute to COVID-19 pathogenesis. Altogether this gives insights into the intricate networks regulating NK cell activation in COVID-19 as well as an outlook towards the role of NK cells in disease pathogenesis.

Overall, we observed a robust NK cell response towards SARS-CoV-2 infection and specific features unique to severe COVID-19 and hyperinflammation, providing a base for understanding the role of NK cells in patients with COVID-19.

## Materials and Methods

### Patient characteristics

SARS-CoV-2 RNA+ patients with moderate or severe COVID-19 disease admitted to the Karolinska University Hospital, Stockholm, Sweden, were recruited to the study. COVID-19 patients were sampled 5-24 days after symptom debut and 0-8 days after being admitted to hospital. Patients classified as having moderate COVID-19 disease had oxygen saturation of 90-94% or were receiving 0.5-3 L/min of oxygen at inclusion. Patients with severe COVID-19 disease were treated in an intensive care unit or a high dependency unit, requiring either non-invasive or mechanical ventilation. Included patients were between 18 and 78 years old. For both groups, patients with current malignant disease and/or ongoing immunomodulatory treatment prior to hospitalization were excluded. The patients were further described by the Sequential Organ Failure Assessment (SOFA) score^53^ and the National Institute of Health (NIH) ordinal scale^54^. Healthy controls were SARS-CoV-2 IgG seronegative at time of inclusion, median age was 50-59 years, and 11 out of 17 were male (65%). The study was approved by the Swedish Ethical Review Authority and all patients gave informed consent. For detailed clinical information, see Supplementary Tables 1 and 2.

### Cell preparation and flow cytometry

Venous blood samples were collected in heparin tubes and peripheral blood mononuclear cells (PBMC) isolated using Ficoll gradient centrifugation. PBMC were thereafter stained fresh with the antibody mix (for antibodies see Supplementary Table 3). Live/Dead cell discrimination was performed using fixable viability dyes (Invitrogen). Cells were permeabilized with the Foxp3/Transcription Factor Staining kit (eBioscience). After staining, cells were fixed with 1% paraformaldehyde for two hours before being acquired on a BD LSR Symphony with 355 nm, 405 nm, 488 nm, 561 nm, and 640 nm lasers.

### Flow cytometry data analysis

FCS3.0 files were exported from the FACSDiva and imported into FlowJo v.10.6.2 for subsequent analysis. The following plugins were used: FlowAI (2.1), DownSample (3.2), UMAP (3.1), PhenoGraph (2.4). First, the data was pre-processed using FlowAI (all checks, second fraction FR = 0.1, alpha FR = 0.01, maximum changepoints = 3, changepoint penalty = 500, dynamic range check side = both) to remove any anomalies present in FCS files. Then, compensation matrix for the 28-color flow cytometry panel was generated using AutoSpill^55^ and applied to files. Dataset as such was used for the downstream analysis in both manual gating and automated analysis. For the automated analysis, events were first downsampled from the NK gate across all samples using DownSample (Supplementary figure 5A and B). Clinical parameter categorical values for each sample were added to downsampled populations as metadata in order to enable identification of these groups, and these were then concatenated for analysis. UMAP was run using all parameters from the panel except BV510 (Live/dead, CD14, CD15, CD19) and PE-Cy5 (CD3). PhenoGraph was run using same parameters from the panel as UMAP (and k = 30). 15.000 cells/sample were exported from the NK gate, apart from six patient samples with fewer events where all cells were taken. When assigning categorical groups formed by different clinical parameters there was an uneven number of patients represented in each group (e.g. 17 ‘healthy controls’, 12 ‘viremic’ and 12 ‘non-viremic’ patients). Over- and under-represented input groups will be similarly weighted in the PhenoGraph output clusters. Therefore, we normalized the PhenoGraph output clusters to account for the total number of cells from each input group. Certain figures were generated in R (versions 3.6.0 and 3.6.1) with packages factoextra (v1.0.5), RColorBrewer (v1.1-2), ggplot2 (v3.2.1 and v3.3.0), tidyr (v.1.0.2), randomcoloR (v.1.1.0.1), reshape2 (v.1.4.3), viridis (v.0.5.1), and pheatmap (v.10.12).

### KIR-ligand genotyping

The DNeasy Blood & Tissue Kit (Qiagen) was used to extract genomic DNA from control and patient whole blood (collected in the EDTA blood tubes). DNA was extracted from the 100 μl of whole blood using the manufacturer’s instructions. DNA was precipitated from the eluate obtained after the last step of the DNeasy Blood & Tissue Kit protocol, with 2.5x volume 100% ethanol and 0.1x volume 3M sodium acetate per reaction and washed with 70% ethanol. DNA concentration was determined with Qubit 4. KIR-ligand genotyping was performed using the KIR HLA ligand kit (Olerup-SSP/CareDx) as per the manufacturer’s instructions.

### Serum protein quantification using PEA

Sera from all patients were evaluated for soluble factors using proximity extension assay technology (PEA, OLINK AB, Uppsala) where 276 selected soluble factors were analyzed.

### Strategy to identify adaptive NK cell expansions

CMV seropositive healthy controls and COVID-19 patients displaying more than 5% of NKG2C+CD57+ cells within their CD56dim NK cell population were considered to have adaptive NK cell expansions (Supplementary Fig. 5a). In all individuals with adaptive expansions, adaptive NK cells displayed higher (> 20%) frequencies of either CD57, CD38, or single KIRs compared to the non-adaptive NK cells (and also in one case high NKG2A). One patient displayed a frequency of adaptive NK cells lower than 5% (3.64%) but was included in the expansion group as co-expression of the other phenotypical markers (differential NKG2A, CD57, CD38 and KIR expression) was in line with what has been described for adaptive NK cells.

### scRNA-seq data analysis

Pre-processed scRNA-seq data was obtained from a published dataset^24^. Data in h5 format was read using Seurat (v3.1.5) then filtered for zero-variance genes and size-factor normalized using scater v1.12.2/scran v1.12.1. CD8 T cells were excluded from downstream NK cell analyses based on k-means clustering of CD8 and NK cell markers from the human cell atlas PBMC benchmarking dataset^56^ followed by re-normalization of the data. Top 1000 highly variable genes were identified using scran (ranked by biological variance and FDR) and used for UMAP dimensionality reduction. Pairwise differential expression of genes detected in at least 20% of cells was performed using MAST (v1.10.0) and genes with an FDR < 10^−3^ in any comparison were clustered into six clusters (determined by gap statistic) by k-means clustering of Z-scores. Gene ontology enrichment of gene clusters was performed using PANTHER overrepresentation tests (release 20200407) using gene ontology database 2020-03-23. Gene set enrichment for bone marrow and blood NK cell signature gene sets^57^ was performed using MAST against a bootstrapped (n = 100) control model of the data and test statistics were combined using the Stouffer method. Data was visualized using ComplexHeatmap (v2.0.0) and ggplot2 (v3.2.1).

### Network analysis

For network analyses, literature-curated interactions from International Molecular Exchange Consortium (IMEx) interactom database, Innate DB, was used^58^. Generic protein-protein interaction network was performed using steiner forest network and visualized using NetworkAnalyst3.0^59^.

### Statistical analysis

Data was analyzed in GraphPad Prism v8. For comparisons between three groups, one-way ANOVA and Kruskal-Wallis test followed by Dunn’s multiple comparisons test was used. For two groups, either parametric or non-parametric matched or non-matched tests were performed. Where indicated, z-score of either median fluorescence intensity (MFI) or frequency of marker expression was calculated as follows: 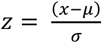, being *x* = raw score, *µ* = mean of sample distribution and *σ* = standard deviation. For categorical comparisons, Fisher’s exact test was used. Significant PhenoGraph clusters (P ≤ 0.05) were determined by Chi-Square goodness-of-fit tests comparing the relative abundance of each categorical group in each individual PhenoGraph cluster relative to input. More details on the exact statistical tests used are mentioned in the respective text/figure legends.

## Data Availability

Curated flow cytometry data is available for exploration via the Karolinska COVID-19 Immune Atlas (homepage currently under construction).

## List of Supplementary Materials

Supplementary Figure 1: NK cell differentiation in COVID-19 disease

Supplementary Figure 2: NK cell activation in COVID-19 disease

Supplementary Figure 3: KIRs and NK cell education in COVID-19 disease

Supplementary Figure 4: Adaptive NK cell expansions in COVID-19

Supplementary Figure 5: Strategy for UMAP analysis and representative marker expression

Supplementary Figure 6: Selected PhenoGraph clusters and their markers are expressed differentially across clinical parameter-defined patient groups

Supplementary Figure 7: Correlations between CD56bright NK cell arming, NK cell phenotype, and soluble factors in COVID-19

Supplementary Table 1: Clinical characteristics of Covid-19 patients

Supplementary Table 2: Clinical laboratory results of Covid-19 patients

Supplementary Table 3: Flow cytometry panel

Supplementary Table 4: Gene ontology analysis of DEGs from scRNAseq analysis of BAL NK cells in COVID-19

Supplementary Table 5: KIR-ligand typing of the study cohort

Supplementary Table 6A: Clinical laboratory results of all patients with and without adaptive NK cell expansions

Supplementary Table 6B: Clinical laboratory results of severe patients with and without adaptive NK cell expansions

Supplementary Table 7: Analysis of the observed distribution of PhenoGraph clusters across clinical parameter-defined groups

Supplementary Table 8A: Literature-curated interactions from International Molecular Exchange Consortium (IMEx) interactom database

Supplementary Table 8B: Nodes and related degrees and betweenness

Supplementary Table 8C: Overview of the KEGG pathways

## Acknowledgements

We thank Craig Hull for methodological discussions and Enrico Lugli’s group for technical assistance with R Studio. This work was supported by the Swedish Research Council, the Swedish Cancer Society, the Swedish Foundation for Strategic Research, Knut and Alice Wallenberg Foundation, Nordstjernan AB, the Center for Innovative Medicine at Karolinska Institutet, Region Stockholm, SRP Diabetes Karolinska Institutet, StratRegen Karolinska Institutet, and Karolinska Institutet. IF and CM are funded by the Wenner-Gren Foundations and QH is supported by the European Molecular Biology Organization (EMBO ALTF 802-2018).

## Author contributions

Conceptualization, C.M., I.F., A.P., N.K.B, Karolinska COVID-19 Study Group; Methodology, C.M., I.F., A.P., Q.H., N.M., D.B., J.M., R.M.H.; Formal analysis, C.M., I.F., A.P., M.C., A.L., Q.H.; Investigation, M.C., B.S., D.B., A.C.G. J.M., N.M., K.S., Resources: S.A., B.R., E.H.A., L.H., A.H.I., M.S., J.K., E.F., M.B., J.K.S., L.I.E., O.R., H.G.L., K.J.M.; Writing – Original Draft, N.K.B; Writing – Review and Editing, All authors; Supervision, N.K.B.

---

**Supplementary Figure 1.**
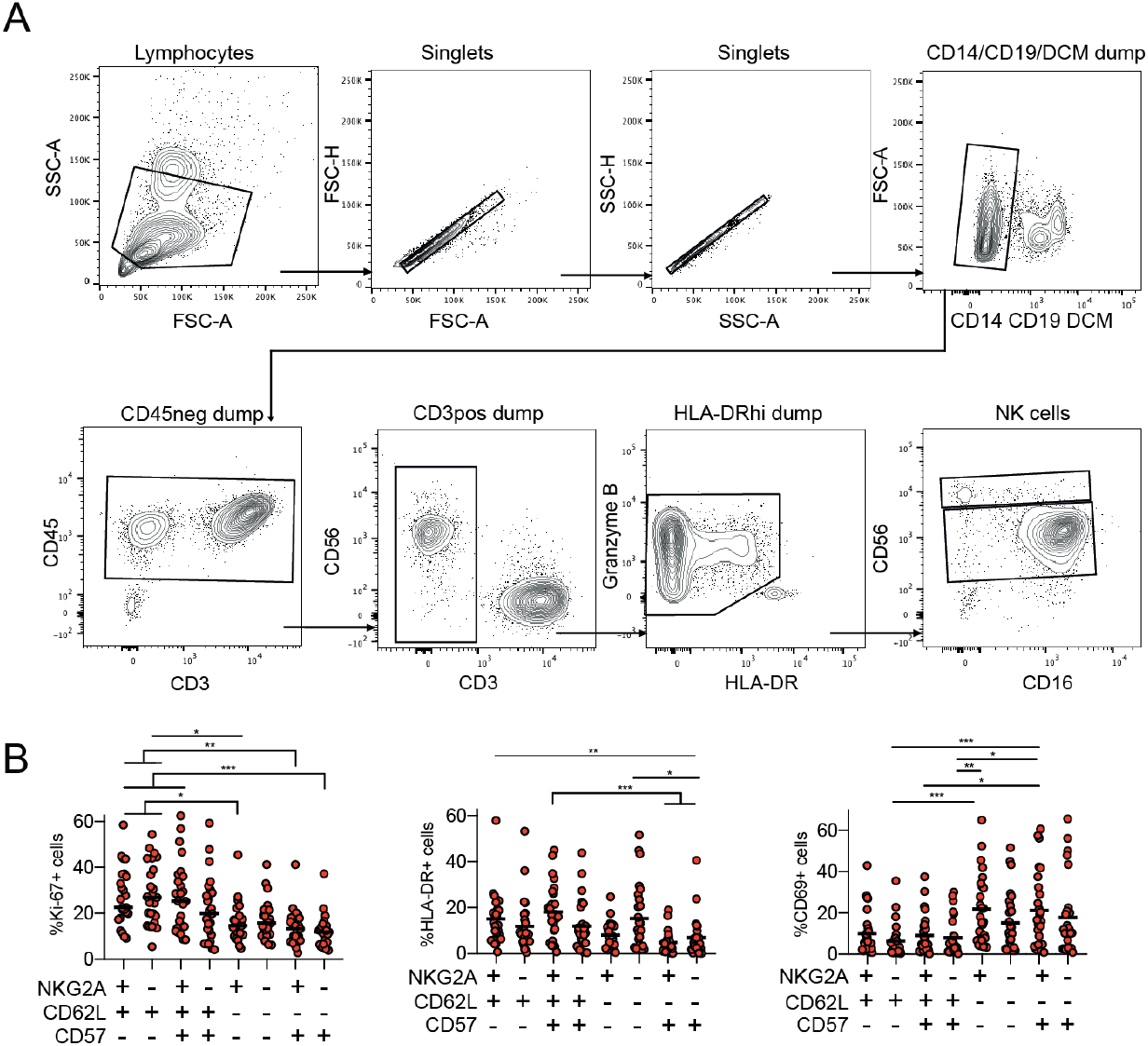
NK cell differentiation in COVID-19 disease. (A) Gating scheme used to identify NK cells as well as CD56bright and CD56dim NK cell subsets. (B) Summary data for expression of Ki67, HLA-DR, and CD69 within CD56dim NK cell subsets defined by expression or not of NKG2A, CD62L, and CD57 in moderate and severe COVID-19 patients (n=25-26). Friedman test was used for statistical comparison, bars represent median, * p < 0.05, ** p < 0.01, ***p < 0.001.

**Supplementary Figure 2.**
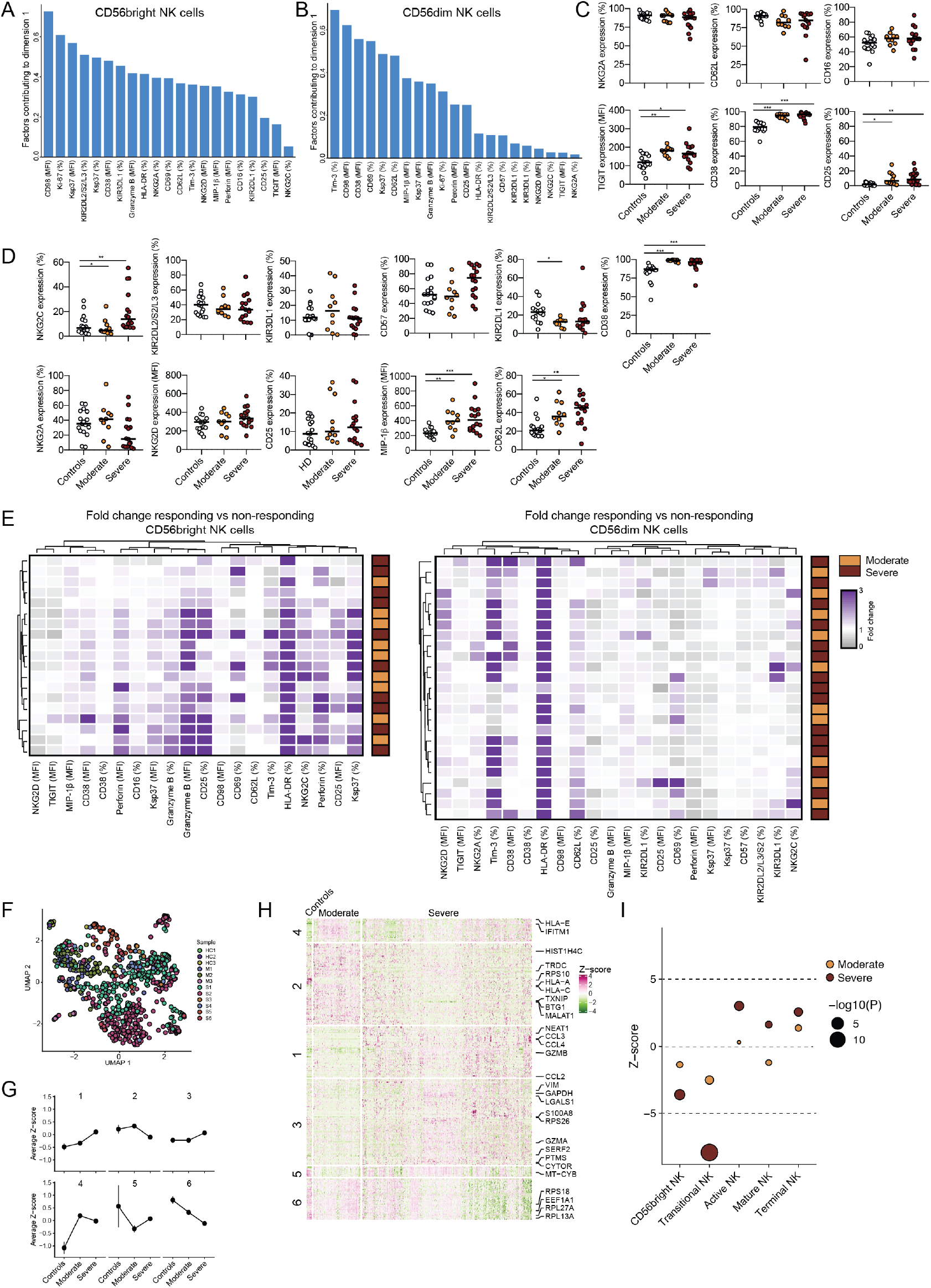
NK cell activation in COVID-19 disease. (A and B) Bar plots showing the parameters that contributed to dimension 1 (dim 1) in Figure 2B PCA plots of (A) CD56bright and (B) CD56dim NK cells. (C) Summary data for the expression of the indicated markers in CD56bright NK cells from healthy controls (n=17), moderate (n=10), and severe (n=15) COVID-19 patients. (D) Summary data for the expression of the indicated markers in CD56dim NK cells from healthy controls (n=17), moderate (n=10), and severe (n=16) COVID-19 patients. (E) Heatmap showing the fold change in expression between Ki-67+ and Ki-67-CD56bright and CD56dim NK cells for the indicated markers after hierarchical clustering in moderate and severe COVID-19 patients. (F) UMAP of scRNAseq data of lung NK cells showing each individual sample that was part of the scRNAseq analysis, HC=healthy control, M=moderate, and S=severe COVID-19 patient. (G) Average Z-score in controls, moderate, and severe COVID-19 patients for the six clusters identified after gene ontology enrichment analysis of genes differentially expressed between controls and patients. (H) Clustered heatmap showing expression of all differentially expressed genes comparing controls and moderate and severe COVID-19 patients. (I) Z-score of indicated NK cell gene sets (obtained from scRNAseq of peripheral blood NK cells) after gene set enrichment analysis. Size of dots indicates significance level using the Stouffer method compared to healthy controls. In C and D, statistical differences were tested using Kruskal-Wallis test followed by Dunn’s multiple comparisons test, bars represent median, *p < 0.05, ** p < 0.01, *** p < 0.001.

**Supplementary Figure 3.**
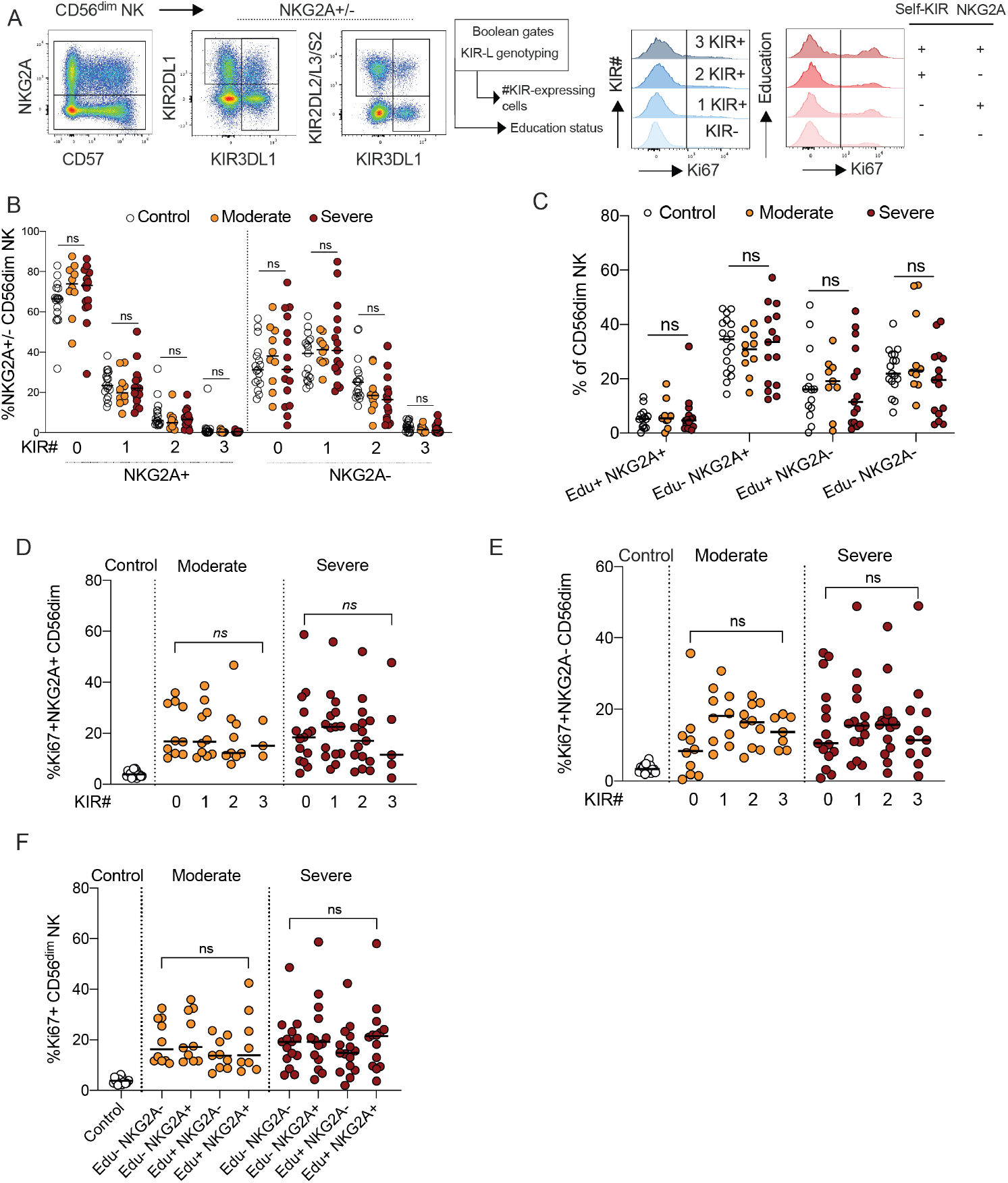
KIRs and NK cell education in COVID-19 disease. (A) Representative flow cytometry plots for NKG2A, KIR2DL1, KIR2DL2/L3/S2, and KIR3DL1 expression on CD56dim NK cells and analysis strategy to identify non-educated and educated CD56dim NK cell subsets as well as subsets expressing different numbers of KIRs. (B) Summary data for the frequency of NKG2A+ or NKG2A-CD56dim NK cells expressing 0, 1, 2, or 3 KIRs in healthy controls (n=17), moderate (n=10), and severe (n=15) COVID-19 patients. (C) Summary data for the frequency of CD56dim NK cell subsets being educated or not via either NKG2A or KIRs (Edu) in healthy controls (n=17 for Edu-, n=14 for Edu+), moderate (n=10 for Edu-, n=9 for Edu+), and severe (n=15) COVID-19 patients. (D-E) Summary data for the frequency of Ki-67+ CD56dim NK cells in healthy controls (n=17), moderate (n=10 for 0/1 KIRs, n=9 for 2 KIRs, n=3 for 3 KIRs), and severe (n=15 for 0/1/2 KIRs, n=5 for 3 KIRs) COVID-19 patients in relation to (D-E) KIR-expression and (F) education status. In D-F, statistical differences were tested using Kruskal-Wallis test followed by Dunn’s multiple comparisons test, bars represent median. Ns, not significant.

**Supplementary Figure 4.**
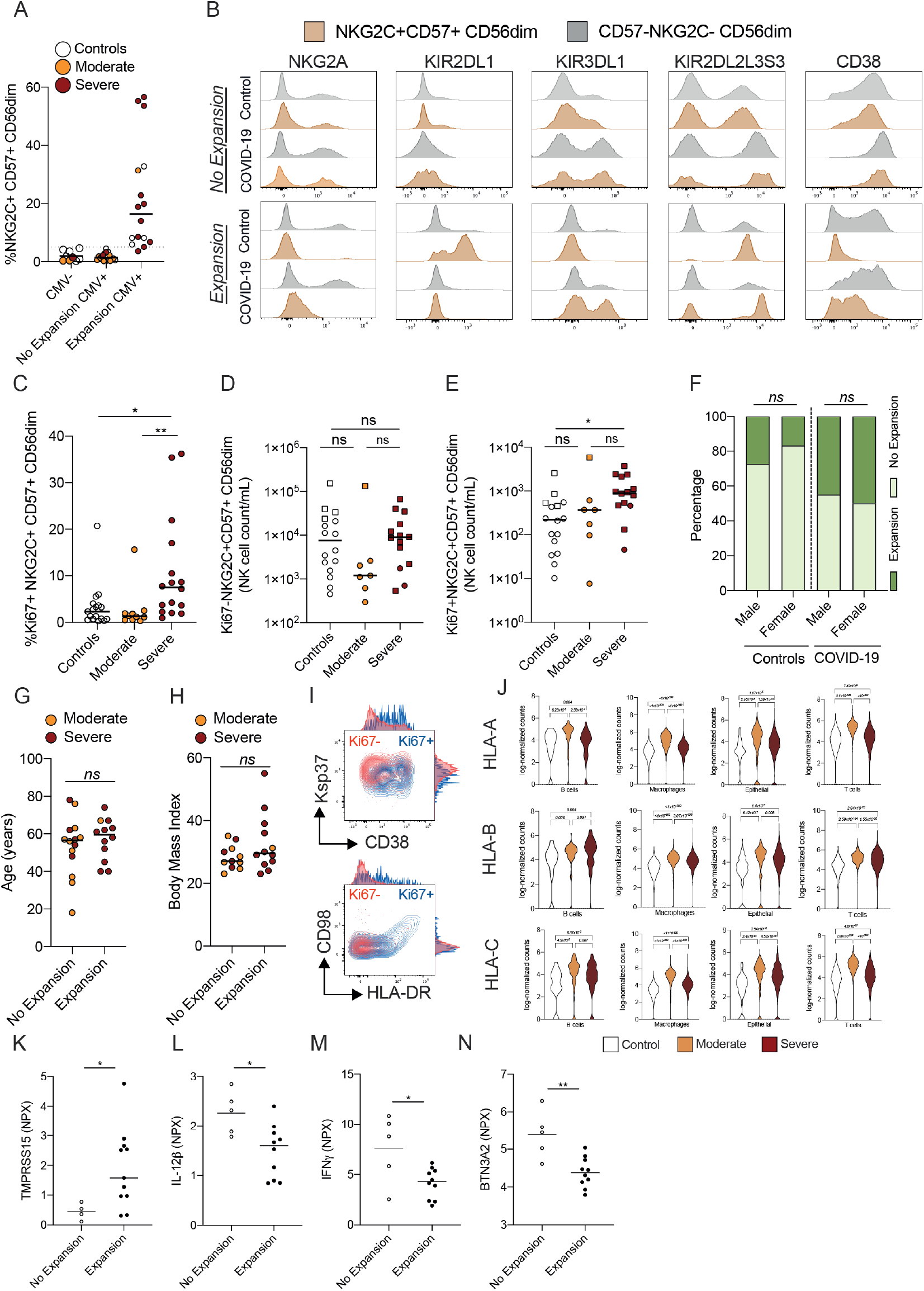
Adaptive NK cell expansions in COVID-19. (A) Summary data for frequency of NKG2C+CD57+ CD56dim NK cells in the indicated groups. (B) Histograms showing representative stainings for NKG2A, KIR2DL1, KIR3DL1, KIR2DL2/L3/S2, and CD38 in NKG2C+CD57+ and NKG2C-CD57-CD56dim NK cells in healthy controls and COVID-19 patients with and without adaptive NK cell expansions. (C-E) Summary data for frequency and absolute count of the indicated subsets in healthy controls (n=17), moderate (n=10), and severe (n=16) COVID-19 patients. Squares represent individuals with NK cell adaptive expansions while circles represent those without expansions. (F) Fraction of individuals from the indicated groups having or not having adaptive NK cell expansions. (G-H) Summary data for age and BMI in COVID-19 patients with and without adaptive NK cell expansions. (I) Representative flow cytometry plots and histograms for the indicated markers in responding (Ki67+, blue) and non-responding (Ki67-, red) adaptive NK cell expansions from one COVID-19 patient. (J) Violin plots showing gene expression of HLA-A, B, and C from scRNAseq data of the indicated lung immune and non-immune cell types in healthy controls, moderate, and severe COVID-19 patients. FDR from pairwise comparisons (see Methods) are shown for statistical significance. In C-E, statistical differences were tested using Kruskal-Wallis test followed by Dunn’s multiple comparisons test, in f using Fisher’s exact test, in G-H using Mann-Whitney test. Bars represent median, ns, not significant, *p < 0.05, ** p < 0.01.

**Supplementary Figure 5.**
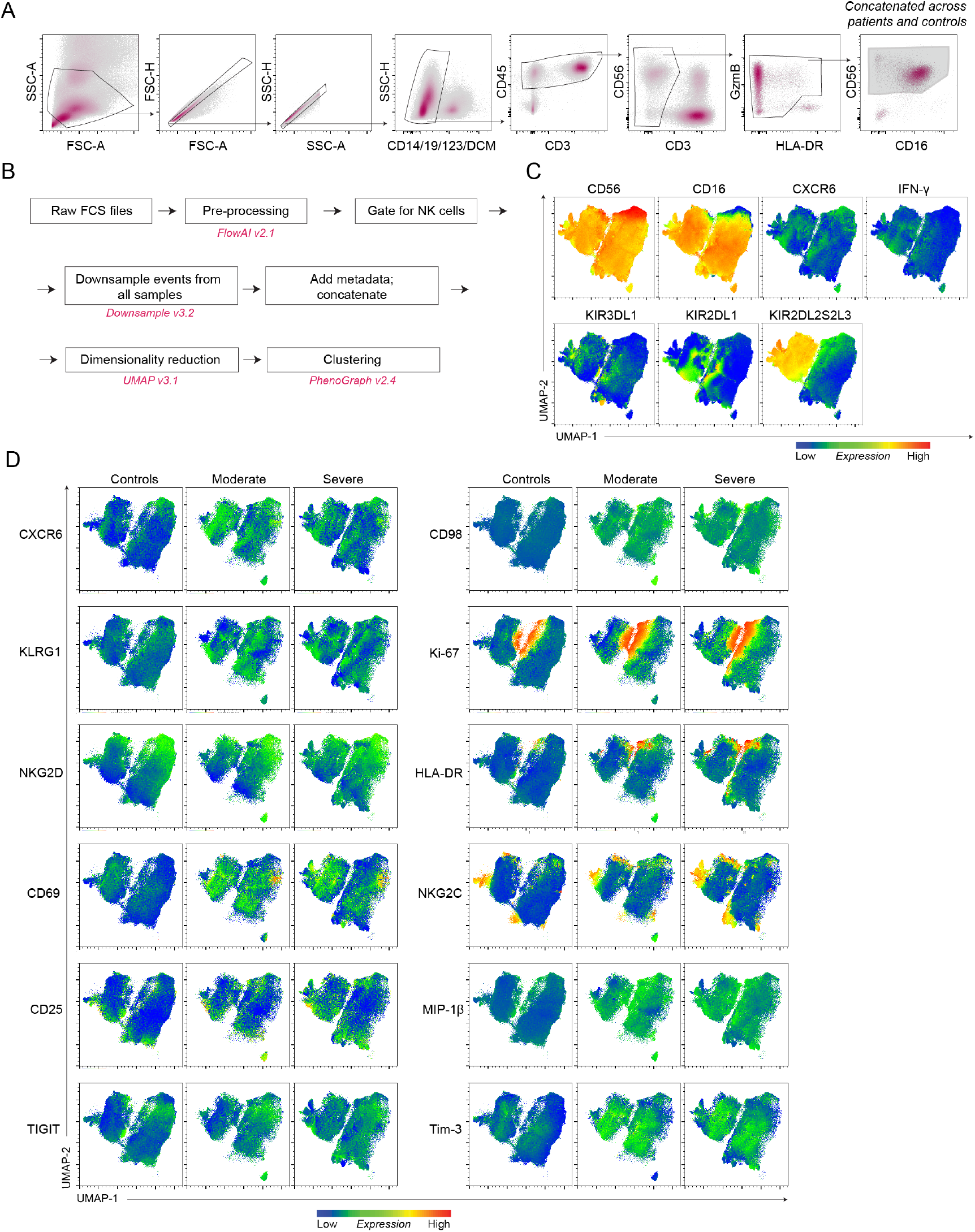
Strategy for UMAP analysis and representative marker expression. (A) Gating scheme used to identify NK cells and subsequently export cells for UMAP analysis. (B) Flow chart of the automated data analysis (Materials and Methods). (C) Representative expression of all other markers except for those shown in Figure 4b included in the dimensionality reduction and clustering analysis. (D) Selected markers of interest (activation, proliferation, inhibitory receptors) shown for controls and moderate and severe COVID-19 patients separately.

**Supplementary Figure 6.**
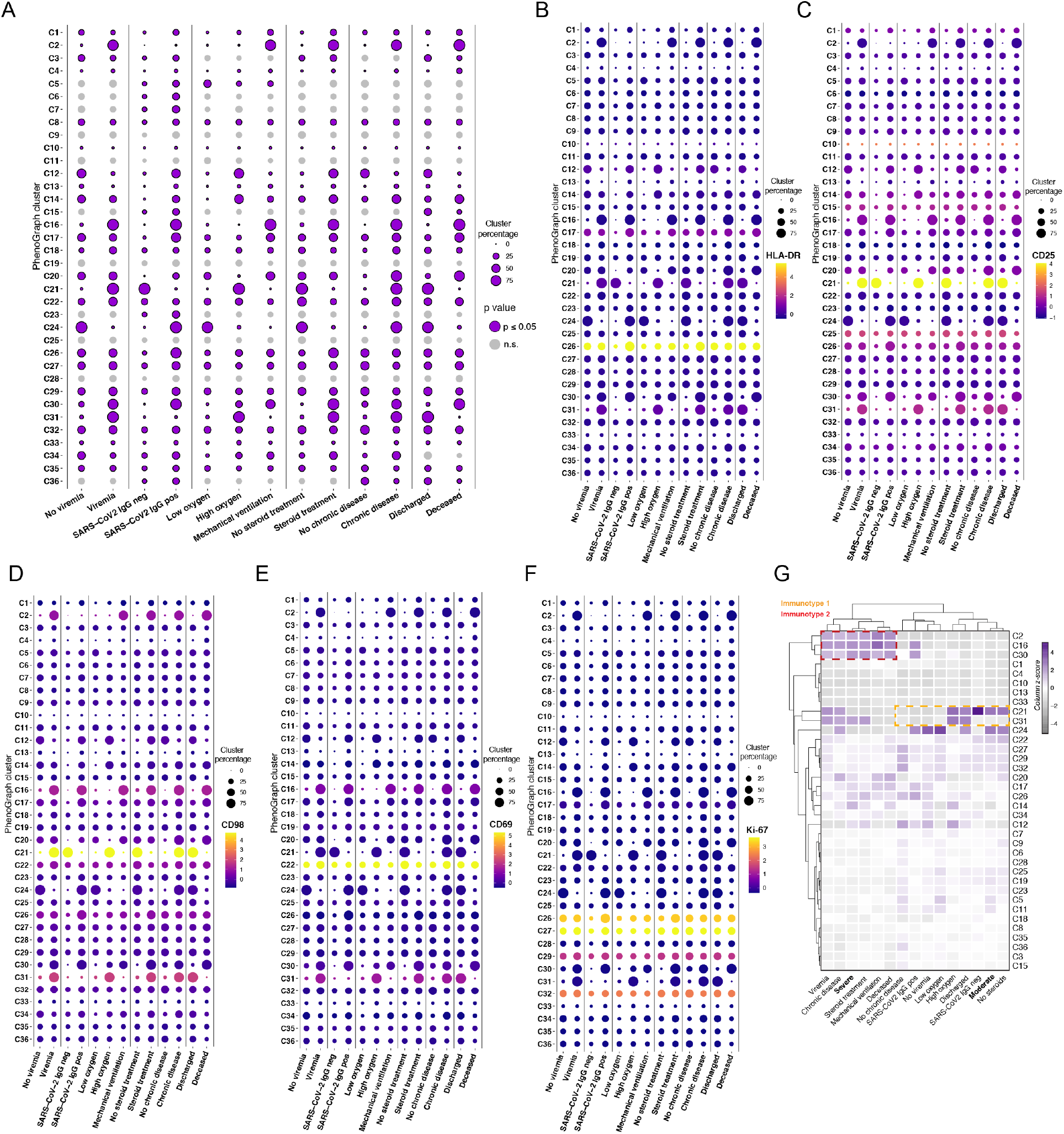
Selected PhenoGraph clusters and their markers are expressed differentially across clinical parameter-defined patient groups. (A) Relative abundance of all 36 PhenoGraph clusters across different patient groups stratified based on the clinical parameter. Only percentages of the PhenoGraph clusters stratifieed within patients are shown (healthy controls are excluded from this graph). Vertical lines indicate separate clinical parameter groups and thus separate comparisons. The size of the circles indicates the size of the cluster. Purple circles with a border indicate significant clusters (p ≤ 0.05) and light-grey circles without a border indicate non-significant clusters (p > 0.05). P values are in Supplementary Table 7 and resulted from the Chi-square goodness-of-fit test (Materials and Methods). (B-F) Median expression of selected markers across all PhenoGraph clusters. Median expression z-score of (B) Ki-67, (C) CD25, (D) CD98, (E) CD69, (F) HLA-DR in all PhenoGraph clusters across different patient groups, stratified based on the clinical parameter. (G) Hierarchical clustering of PhenoGraph clusters and clinical categorical parameters. Heatmap calculated as a column z-score of cluster percentages, including relative abundance of clusters in moderate and severe groups of patients. Putative NK immunotypes (Immunotype 1 and Immunotype 2) are indicated.

**Supplementary Figure 7.**
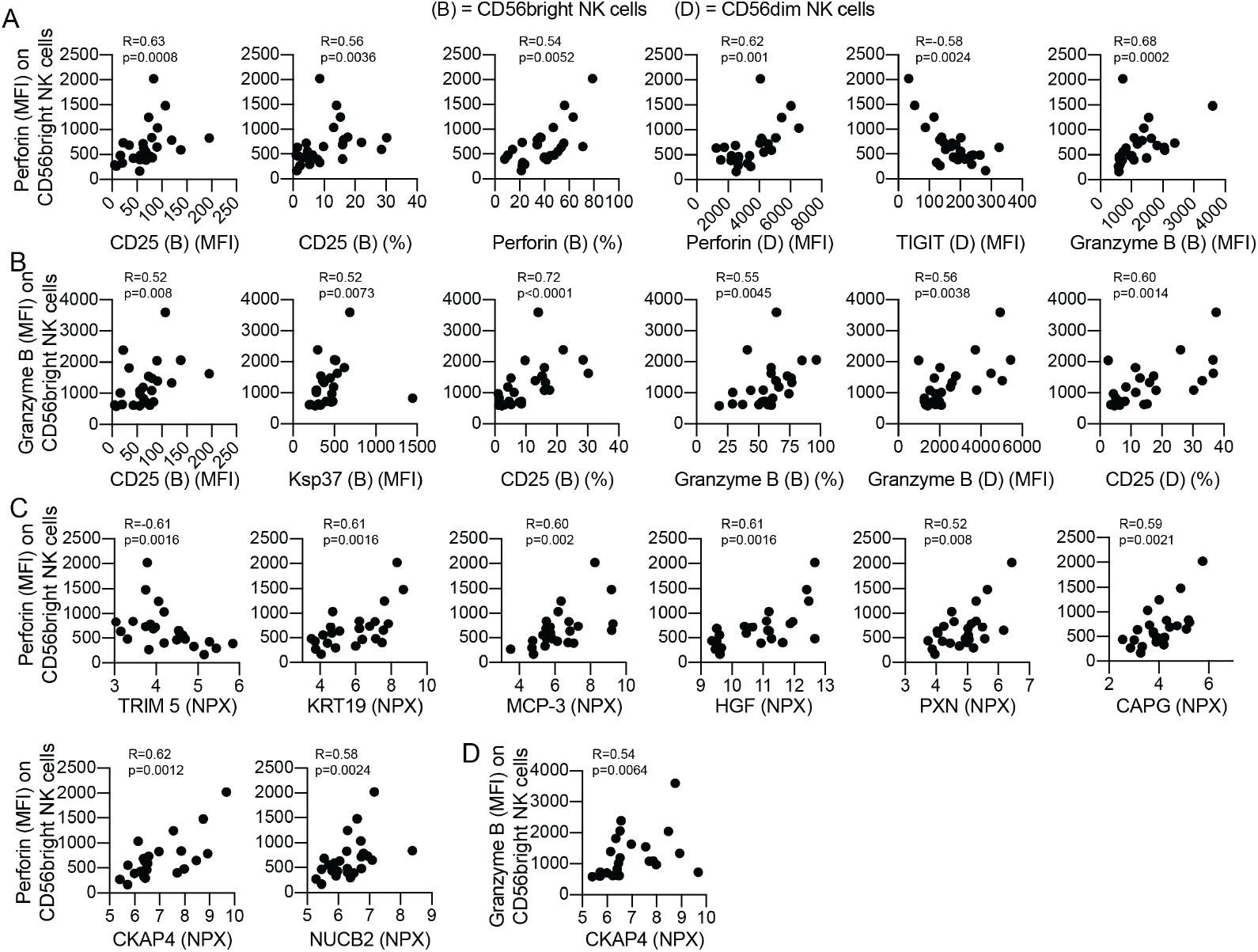
Correlations between CD56bright NK cell arming, NK cell phenotype, and soluble factors in COVID-19. (A) Correlation plots between perforin (MFI) expression on CD56bright NK cells and the indicated markers. (B) Correlation plots between granzyme B (MFI) expression on CD56bright NK cells and the indicated markers. (C) Correlation plots between perforin or granzyme B (both MFI) expression on CD56bright NK cells and the indicated soluble factors. For A-C, n=24-25, Spearman correlations were performed, and R and p-values are indicated in each figure. In A and B, “(B)” denotes correlation against an NK cell parameter within CD56bright NK cells and “(D)” within CD56dim NK cells.

**Supplementary table 1.**
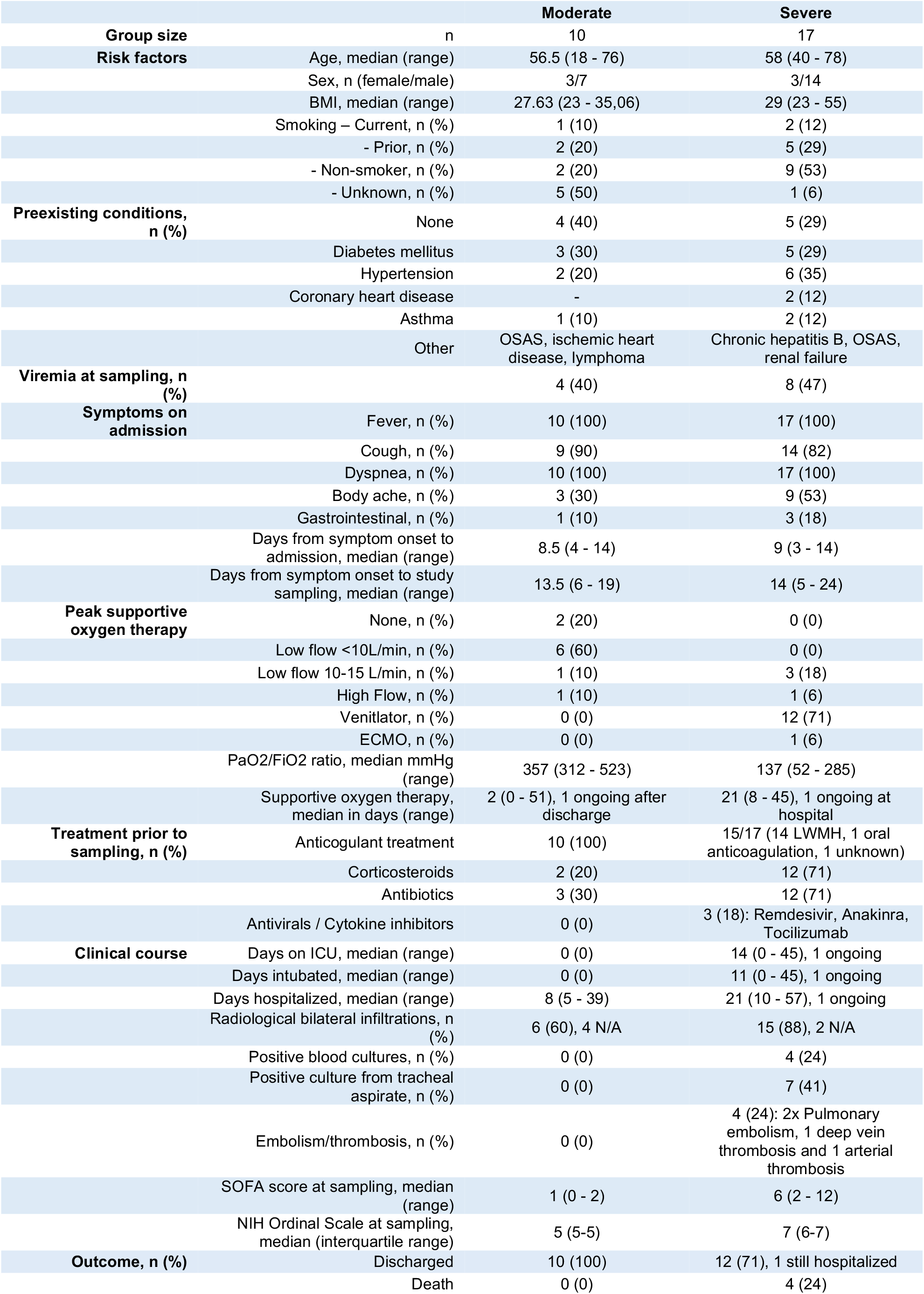
Clinical characteristics of Covid-19 patients.

**Supplementary Table 2.** Clinical laboratory results of Covid-19 patients.

|  | Normal range | Moderate | Severe/Critical | p-value |
| --- | --- | --- | --- | --- |
| **Before study sampling** |  |  |  |  |
|  |  | *Median peak (range)* | *Median peak (range)* |  |
| C-reactive protein, mg/L | <3 | 148 (34 - 346) | 296 (101 - 452) | 0.015 |
| Procalcitonin, µg/L | <0.5 | 0.28 (0.11 - 66) | 0.90 (0.19 - 10) | 0.10 |
| D-dimer, mg/L | <0.56 | 0.72 (0.28 - 1.5) | 2.3 (0.58 - 5.4) | <0.001 |
| Ferritin, µg/L | 10-150 | 856 (219 - 3570) | 1793 (183 - 5500) | 0.074 |
| Fibrinogen, g/L | 2-4.2 | N/A | 7.05 (4.1 - 10.2) |  |
| Lactate dehydrogenase, µkat/L | <3.5 | 5.6 (3.8 - 13.7) | 10.6 (0.89 - 23) | 0.003 |
| Alanine aminotransferase, µkat/L | <0.76 | 0.67 (0.51 - 3.43) | 1.24 (0.51 - 13.8) | 0.16 |
| Bilirubin, µmol/L | <26 | 8 (4 - 11) | 12 (7 - 81) | 0.002 |
| Creatinine, µmol/L | <90 | 75 (49 - 168) | 90 (46 - 326) | 0.20 |
| Prothrombin complex, INR | <1.2 | 1.0 (0.9 - 1.1) | 1.1 (0.9 - 2.1) | 0.005 |
| Troponin T, ng/L | <15 | 11 (5 - 218) | 19 (8 - 601) | 0.037 |
| Myoglobin, µg/L | <73 | 23 (22 - 37) | 369 (36 - 6400) | 0.001 |
| Platelet count, highest, x10<sup>9</sup>/L | 165-387 | 296 (152 - 642) | 398 (149 - 770) | 0.31 |
| Platelet count, lowest, x10<sup>9</sup>/L | 165-387 | 232 (131 - 342) | 196 (110 - 330) | 0.86 |
| Lymphocytes, lowest, x10<sup>9</sup>/L | 1.1-3.5 | 0.8 (0.3 - 1.9) | 0.6 (0.2 - 1.5) | 0.040 |
| Neutrophils, lowest, x10<sup>9</sup>/L | 1.6-5.9 | 4.0 (0.5 - 6.8) | 4.4 (1.7 - 15.9) | 0.13 |
| **At study sampling (+/-24h)** |  |  |  |  |
| C-reactive protein, mg/L | <3 | 104 (20 - 394) | 203 (37 - 346) | 0.11 |
| Procalcitonin, µg/L | <0.5 | 0.39 (0.12 - 866) | 0.52 (0.16 - 10) | 0.59 |
| Leukocytes, highest, x 10<sup>9</sup>/L | 3.5-8.8 | 7.7 (1.5 - 12.6) | 12.5 (4 - 18.4) | 0.005 |
| Leukocytes, lowest, x 10<sup>9</sup>/L | 3.5-8.8 | 6.2 (1 - 10.5) | 9.2 (2.3 - 15.7) | 0.013 |
| Neutrophils, highest, x 10<sup>9</sup>/L | 1.6-5.9L | 5.4 (0.9 - 9.3) | 10.9 (3.2 - 16.7) | 0.001 |
| D-dimer, mg/L | <0.56 | 0.71 (0.36 - 1.3) | 2.3 (0.44 - 7.6) | 0.001 |
| Ferritin, µg/L | 10-150 | 738 (161 - 1971) | 1832 (183 - 4306) | 0.053 |
| Fibrinogen, g/L | 2-4.2 | N/A | 6.4 (3.5 - 10.2) |  |
| Bilirubin, µmol/L | <26 | 6 (4 - 11) | 9 (6 - 81) | 0.049 |
| Creatinine, µmol/L | <90 | 68 (43 - 168) | 73 (36 - 326) | 0.47 |
| Prothrombin complex, INR | <1.2 | 1.0 (1 - 1.1) | 1.1 (0.9 - 1.4) | 0.32 |
| Troponin T, ng/L | <15 | 6 (5 - 43) | 18 (6 - 434) | 0.018 |
| Haemoglobin, lowest, g/L | 117-153 | 124 (90 - 145) | 113 (73 - 146) | 0.19 |
| Lymphocytes, lowest, x 10<sup>9</sup>/L | 1.1-3.5 | 1.2 (0.3 - 2.4) | 0.8 (0.3 - 2.3) | 0.023 |
| Albumin, lowest, g/L | 36-48 | 28 (21 - 37) | 19 (16 - 29) | 0.003 |
| Platelet count, lowest, x 10<sup>9</sup>/L | 165-387 | 324 (131 - 642) | 361 (146 - 579) | 0.49 |
| **At study sampling (+/-5d)** |  |  |  |  |
| Interleukin-6, ng/L | <7 | 58 (2 - 151) | 198 (29 - 8093) | 0.041 |
| Interleukin-1β, ng/L | <5 | N/A | 9.6 (5.4 - 16.8) |  |
| Interleukin-10, ng/L | <5 | N/A | 24 (5.6 - 58) |  |
| Tumor necrosis factor-α, ng/L | <12 | N/A | 15 (8 - 48) |  |

**Supplementary Table 3.**
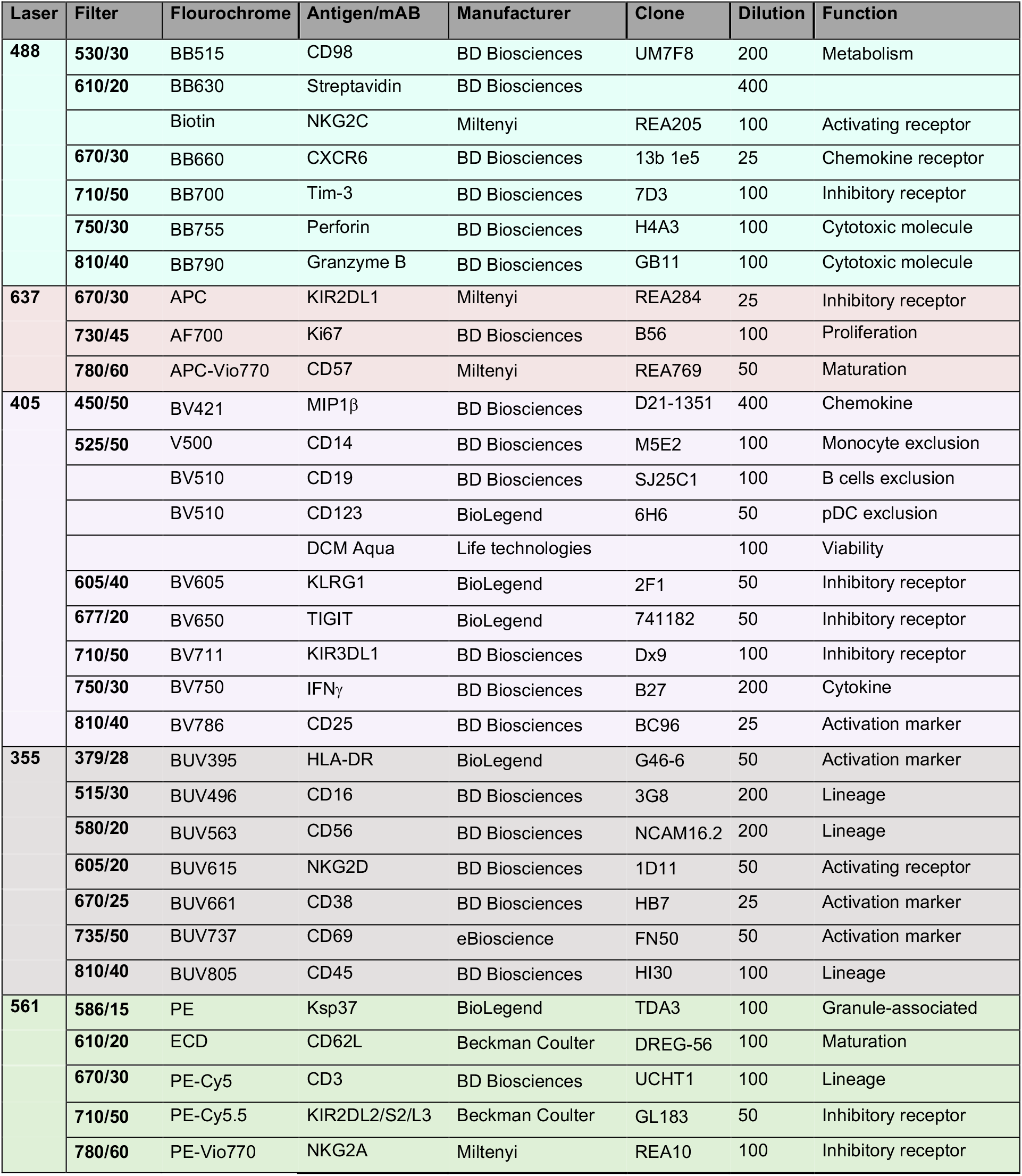
Flow cytometry panel.

| Laser | Filter | Fluorochrome | Antigen/mAB | Manufacturer | Clone | Dilution | Function |
| --- | --- | --- | --- | --- | --- | --- | --- |
| 488 | 530/30 | BB515 | CD98 | BD Biosciences | UM7F8 | 200 | Metabolism |
|  | 610/20 | BB630 | Streptavidin | BD Biosciences |  | 400 |  |
|  |  | Biotin | NKG2C | Miltenyi | REA205 | 100 | Activating receptor |
|  | 670/30 | BB660 | CXCR6 | BD Biosciences | 13b 1e5 | 25 | Chemokine receptor |
|  | 710/50 | BB700 | Tim-3 | BD Biosciences | 7D3 | 100 | Inhibitory receptor |
|  | 750/30 | BB755 | Perforin | BD Biosciences | H4A3 | 100 | Cytotoxic molecule |
|  | 810/40 | BB790 | Granzyme B | BD Biosciences | GB11 | 100 | Cytotoxic molecule |
| 637 | 670/30 | APC | KIR2DL1 | Miltenyi | REA284 | 25 | Inhibitory receptor |
|  | 730/45 | AF700 | Ki67 | BD Biosciences | B56 | 100 | Proliferation |
|  | 780/60 | APC-Vio770 | CD57 | Miltenyi | REA769 | 50 | Maturation |
| 405 | 450/50 | BV421 | MIP1β | BD Biosciences | D21-1351 | 400 | Chemokine |
|  | 525/50 | V500 | CD14 | BD Biosciences | M5E2 | 100 | Monocyte exclusion |
|  |  | BV510 | CD19 | BD Biosciences | SJ25C1 | 100 | B cells exclusion |
|  |  | BV510 | CD123 | BioLegend | 6H6 | 50 | pDC exclusion |
|  |  |  | DCM Aqua | Life technologies |  | 100 | Viability |
|  | 605/40 | BV605 | KLRG1 | BioLegend | 2F1 | 50 | Inhibitory receptor |
|  | 677/20 | BV650 | TIGIT | BioLegend | 741182 | 50 | Inhibitory receptor |
|  | 710/50 | BV711 | KIR3DL1 | BD Biosciences | Dx9 | 100 | Inhibitory receptor |
| | 750/30 | BV750 | IFNγ | BD Biosciences | B27 | 200 | Cytokine |
|  | 810/40 | BV786 | CD25 | BD Biosciences | BC96 | 25 | Activation marker |
| 355 | 379/28 | BUV395 | HLA-DR | BioLegend | G46-6 | 50 | Activation marker |
|  | 515/30 | BUV496 | CD16 | BD Biosciences | 3G8 | 200 | Lineage |
|  | 580/20 | BUV563 | CD56 | BD Biosciences | NCAM16.2 | 200 | Lineage |
|  | 605/20 | BUV615 | NKG2D | BD Biosciences | 1D11 | 50 | Activating receptor |
|  | 670/25 | BUV661 | CD38 | BD Biosciences | HB7 | 25 | Activation marker |
|  | 735/50 | BUV737 | CD69 | eBioscience | FN50 | 50 | Activation marker |
|  | 810/40 | BUV805 | CD45 | BD Biosciences | HI30 | 100 | Lineage |
| 561 | 586/15 | PE | Ksp37 | BioLegend | TDA3 | 100 | Granule-associated |
|  | 610/20 | ECD | CD62L | Beckman Coulter | DREG-56 | 100 | Maturation |
|  | 670/30 | PE-Cy5 | CD3 | BD Biosciences | UCHT1 | 100 | Lineage |
|  | 710/50 | PE-Cy5.5 | KIR2DL2/S2/L3 | Beckman Coulter | GL183 | 50 | Inhibitory receptor |
|  | 780/60 | PE-Vio770 | NKG2A | Miltenyi | REA10 | 100 | Inhibitory receptor |

**Supplementary Table 5.** KIR-ligand typing.

| Sample ID | KIR-ligand genotype |  |
| --- | --- | --- |
|  | *HLA-C locus* | *HLA-B locus* |
| Control 1 | C1/C2 | Bw4/Bw6 |
| Control 2 | C1/C1 | Bw4/Bw6 |
| Control 3 | C1/C2 | Bw6/Bw6 |
| Control 4 | C1/C2 | Bw4/Bw6 |
| Control 5 | C1/C1 | Bw4/Bw4 |
| Control 6 | C1/C1 | Bw4/Bw6 |
| Control 7 | C1/C2 | Bw6/Bw6 |
| Control 8 | C1/C1 | Bw4/Bw6 |
| Control 9 | C2/C2 | Bw4/Bw4 |
| Control 10 | C1/C1 | Bw6/Bw6 |
| Control 11 | C1/C2 | Bw6/Bw6 |
| Control 12 | C1/C1 | Bw6/Bw6 |
| Control 13 | C1/C2 | Bw4/Bw6 |
| Control 14 | C1/C1 | Bw6/Bw6 |
| Control 15 | C1/C2 | Bw6/Bw6 |
| Control 16 | C1/C1 | Bw4/Bw6 |
| Control 17 | C1/C2 | Bw4/Bw6 |
| Patient 1 | C1/C2 | Bw6/Bw6 |
| Patient 2 | C1/C2 | Bw4/Bw4 |
| Patient 3 | C1/C2 | Bw4/Bw4 |
| Patient 4 | C1/C2 | Bw4/Bw6 |
| Patient 5 | C1/C2 | Bw4/Bw4 |
| Patient 6 | C1/C2 | Bw6/Bw6 |
| Patient 7 | C1/C2 | Bw4/Bw6 |
| Patient 8 | C1/C1 | Bw4/Bw4 |
| Patient 9 | C1/C2 | Bw4/Bw6 |
| Patient 10 | C1/C1 | Bw4/Bw6 |
| Patient 11 | C1/C2 | Bw4/Bw6 |
| Patient 12 | C2/C2 | Bw4/Bw6 |
| Patient 13 | C1/C1 | Bw4/Bw6 |
| Patient 14 | C1/C1 | Bw4/Bw6 |
| Patient 15 | C1/C1 | Bw4/Bw6 |
| Patient 16 | C2/C2 | Bw4/Bw6 |
| Patient 17 | C1/C2 | Bw4/Bw6 |
| Patient 18 | C1/C2 | Bw4/Bw6 |
| Patient 19 | C1/C2 | Bw4/Bw6 |
| Patient 20 | C1/C2 | Bw4/Bw6 |
| Patient 21 | C1/C1 | Bw4/Bw4 |
| Patient 22 | C1/C2 | Bw4/Bw4 |
| Patient 23 | C1/C1 | Bw6/Bw6 |
| Patient 24 | C1/C2 | Bw4/Bw6 |
| Patient 25 | C1/C2 | Bw4/Bw6 |
| Patient 26 | C1/C1 | Bw4/Bw6 |
| Patient 27 | C1/C2 | Bw4/Bw6 |

**Supplementary Table 6A.**
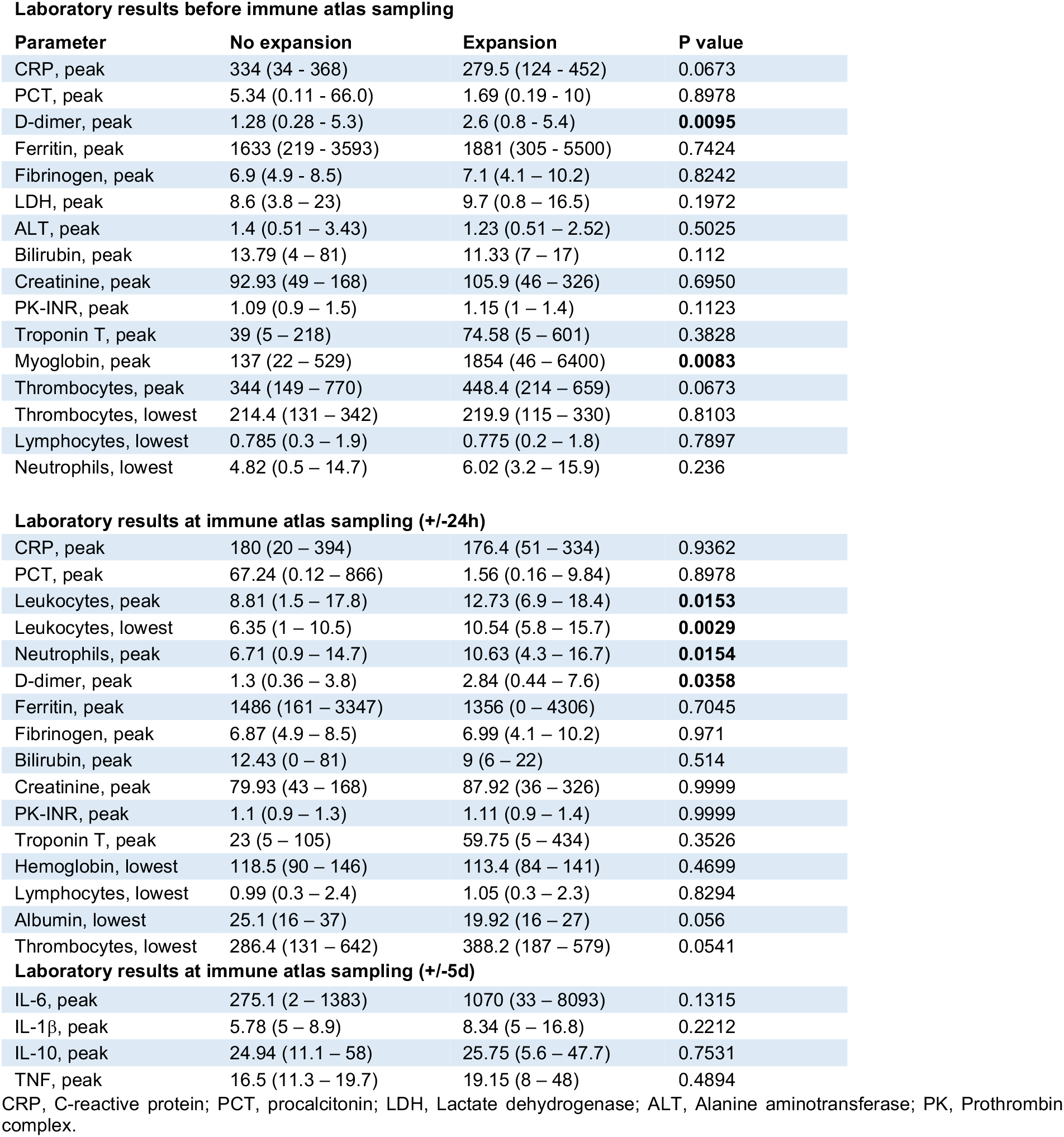
Clinical laboratory results of all patients with and without adaptive NK cell expansions.

Laboratory results before immune atlas sampling
| Parameter | No expansion | Expansion | P value |
| --- | --- | --- | --- |
| CRP, peak | 334 (34 - 368) | 279.5 (124 - 452) | 0.0673 |
| PCT, peak | 5.34 (0.11 - 66.0) | 1.69 (0.19 - 10) | 0.8978 |
| D-dimer, peak | 1.28 (0.28 - 5.3) | 2.6 (0.8 - 5.4) | **0.0095** |
| Ferritin, peak | 1633 (219 - 3593) | 1881 (305 - 5500) | 0.7424 |
| Fibrinogen, peak | 6.9 (4.9 - 8.5) | 7.1 (4.1 - 10.2) | 0.8242 |
| LDH, peak | 8.6 (3.8 - 23) | 9.7 (0.8 - 16.5) | 0.1972 |
| ALT, peak | 1.4 (0.51 - 3.43) | 1.23 (0.51 - 2.52) | 0.5025 |
| Bilirubin, peak | 13.79 (4 - 81) | 11.33 (7 - 17) | 0.112 |
| Creatinine, peak | 92.93 (49 - 168) | 105.9 (46 - 326) | 0.6950 |
| PK-INR, peak | 1.09 (0.9 - 1.5) | 1.15 (1 - 1.4) | 0.1123 |
| Troponin T, peak | 39 (5 - 218) | 74.58 (5 - 601) | 0.3828 |
| Myoglobin, peak | 137 (22 - 529) | 1854 (46 - 6400) | **0.0083** |
| Thrombocytes, peak | 344 (149 - 770) | 448.4 (214 - 659) | 0.0673 |
| Thrombocytes, lowest | 214.4 (131 - 342) | 219.9 (115 - 330) | 0.8103 |
| Lymphocytes, lowest | 0.785 (0.3 - 1.9) | 0.775 (0.2 - 1.8) | 0.7897 |
| Neutrophils, lowest | 4.82 (0.5 - 14.7) | 6.02 (3.2 - 15.9) | 0.236 |
| Laboratory results at immune atlas sampling (+/-24h) |  |  |  |
| CRP, peak | 180 (20 - 394) | 176.4 (51 - 334) | 0.9362 |
| PCT, peak | 67.24 (0.12 - 866) | 1.56 (0.16 - 9.84) | 0.8978 |
| Leukocytes, peak | 8.81 (1.5 - 17.8) | 12.73 (6.9 - 18.4) | **0.0153** |
| Leukocytes, lowest | 6.35 (1 - 10.5) | 10.54 (5.8 - 15.7) | **0.0029** |
| Neutrophils, peak | 6.71 (0.9 - 14.7) | 10.63 (4.3 - 16.7) | **0.0154** |
| D-dimer, peak | 1.3 (0.36 - 3.8) | 2.84 (0.44 - 7.6) | **0.0358** |
| Ferritin, peak | 1486 (161 - 3347) | 1356 (0 - 4306) | 0.7045 |
| Fibrinogen, peak | 6.87 (4.9 - 8.5) | 6.99 (4.1 - 10.2) | 0.971 |
| Bilirubin, peak | 12.43 (0 - 81) | 9 (6 - 22) | 0.514 |
| Creatinine, peak | 79.93 (43 - 168) | 87.92 (36 - 326) | 0.9999 |
| PK-INR, peak | 1.1 (0.9 - 1.3) | 1.11 (0.9 - 1.4) | 0.9999 |
| Troponin T, peak | 23 (5 - 105) | 59.75 (5 - 434) | 0.3526 |
| Hemoglobin, lowest | 118.5 (90 - 146) | 113.4 (84 - 141) | 0.4699 |
| Lymphocytes, lowest | 0.99 (0.3 - 2.4) | 1.05 (0.3 - 2.3) | 0.8294 |
| Albumin, lowest | 25.1 (16 - 37) | 19.92 (16 - 27) | 0.056 |
| Thrombocytes, lowest | 286.4 (131 - 642) | 388.2 (187 - 579) | 0.0541 |
| Laboratory results at immune atlas sampling (+/-5d) |  |  |  |
| IL-6, peak | 275.1 (2 - 1383) | 1070 (33 - 8093) | 0.1315 |
| IL-1β, peak | 5.78 (5 - 8.9) | 8.34 (5 - 16.8) | 0.2212 |
| IL-10, peak | 24.94 (11.1 - 58) | 25.75 (5.6 - 47.7) | 0.7531 |
| TNF, peak | 16.5 (11.3 - 19.7) | 19.15 (8 - 48) | 0.4894 |

CRP, C-reactive protein; PCT, procalcitonin; LDH, Lactate dehydrogenase; ALT, Alanine aminotransferase; PK, Prothrombin complex.

**Supplementary Table 6B.**
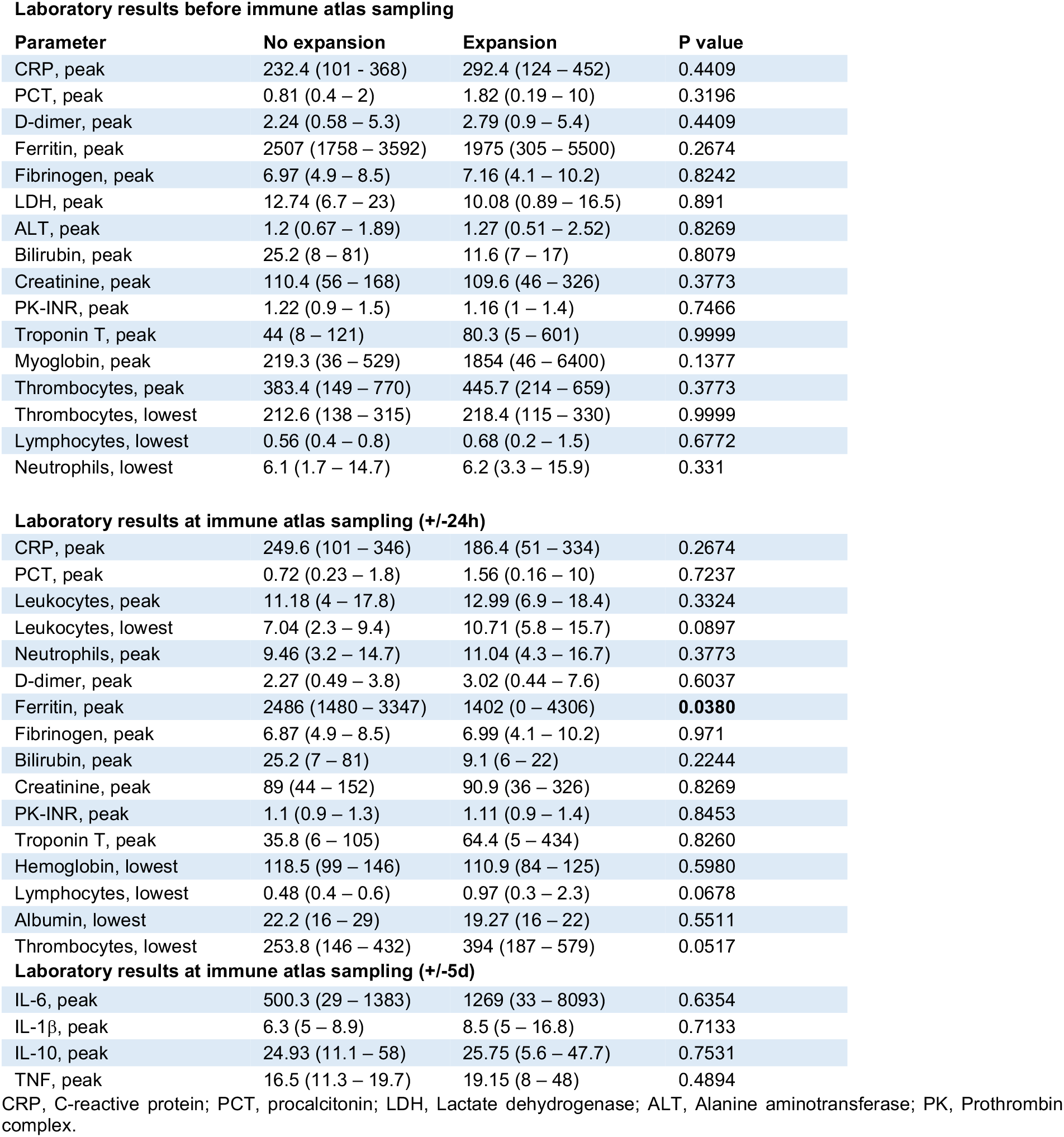
Clinical laboratory results of severe patients with and without adaptive NK cell expansions.

Laboratory results before immune atlas sampling
| Parameter | No expansion | Expansion | P value |
| --- | --- | --- | --- |
| CRP, peak | 232.4 (101 - 368) | 292.4 (124 - 452) | 0.4409 |
| PCT, peak | 0.81 (0.4 - 2) | 1.82 (0.19 - 10) | 0.3196 |
| D-dimer, peak | 2.24 (0.58 - 5.3) | 2.79 (0.9 - 5.4) | 0.4409 |
| Ferritin, peak | 2507 (1758 - 3592) | 1975 (305 - 5500) | 0.2674 |
| Fibrinogen, peak | 6.97 (4.9 - 8.5) | 7.16 (4.1 - 10.2) | 0.8242 |
| LDH, peak | 12.74 (6.7 - 23) | 10.08 (0.89 - 16.5) | 0.891 |
| ALT, peak | 1.2 (0.67 - 1.89) | 1.27 (0.51 - 2.52) | 0.8269 |
| Bilirubin, peak | 25.2 (8 - 81) | 11.6 (7 - 17) | 0.8079 |
| Creatinine, peak | 110.4 (56 - 168) | 109.6 (46 - 326) | 0.3773 |
| PK-INR, peak | 1.22 (0.9 - 1.5) | 1.16 (1 - 1.4) | 0.7466 |
| Troponin T, peak | 44 (8 - 121) | 80.3 (5 - 601) | 0.9999 |
| Myoglobin, peak | 219.3 (36 - 529) | 1854 (46 - 6400) | 0.1377 |
| Thrombocytes, peak | 383.4 (149 - 770) | 445.7 (214 - 659) | 0.3773 |
| Thrombocytes, lowest | 212.6 (138 - 315) | 218.4 (115 - 330) | 0.9999 |
| Lymphocytes, lowest | 0.56 (0.4 - 0.8) | 0.68 (0.2 - 1.5) | 0.6772 |
| Neutrophils, lowest | 6.1 (1.7 - 14.7) | 6.2 (3.3 - 15.9) | 0.331 |
| Laboratory results at immune atlas sampling (+/-24h) |  |  |  |
| CRP, peak | 249.6 (101 - 346) | 186.4 (51 - 334) | 0.2674 |
| PCT, peak | 0.72 (0.23 - 1.8) | 1.56 (0.16 - 10) | 0.7237 |
| Leukocytes, peak | 11.18 (4 - 17.8) | 12.99 (6.9 - 18.4) | 0.3324 |
| Leukocytes, lowest | 7.04 (2.3 - 9.4) | 10.71 (5.8 - 15.7) | 0.0897 |
| Neutrophils, peak | 9.46 (3.2 - 14.7) | 11.04 (4.3 - 16.7) | 0.3773 |
| D-dimer, peak | 2.27 (0.49 - 3.8) | 3.02 (0.44 - 7.6) | 0.6037 |
| Ferritin, peak | 2486 (1480 - 3347) | 1402 (0 - 4306) | **0.0380** |
| Fibrinogen, peak | 6.87 (4.9 - 8.5) | 6.99 (4.1 - 10.2) | 0.971 |
| Bilirubin, peak | 25.2 (7 - 81) | 9.1 (6 - 22) | 0.2244 |
| Creatinine, peak | 89 (44 - 152) | 90.9 (36 - 326) | 0.8269 |
| PK-INR, peak | 1.1 (0.9 - 1.3) | 1.11 (0.9 - 1.4) | 0.8453 |
| Troponin T, peak | 35.8 (6 - 105) | 64.4 (5 - 434) | 0.8260 |
| Hemoglobin, lowest | 118.5 (99 - 146) | 110.9 (84 - 125) | 0.5980 |
| Lymphocytes, lowest | 0.48 (0.4 - 0.6) | 0.97 (0.3 - 2.3) | 0.0678 |
| Albumin, lowest | 22.2 (16 - 29) | 19.27 (16 - 22) | 0.5511 |
| Thrombocytes, lowest | 253.8 (146 - 432) | 394 (187 - 579) | 0.0517 |
| Laboratory results at immune atlas sampling (+/-5d) |  |  |  |
| IL-6, peak | 500.3 (29 - 1383) | 1269 (33 - 8093) | 0.6354 |
| IL-1β, peak | 6.3 (5 - 8.9) | 8.5 (5 - 16.8) | 0.7133 |
| IL-10, peak | 24.93 (11.1 - 58) | 25.75 (5.6 - 47.7) | 0.7531 |
| TNF, peak | 16.5 (11.3 - 19.7) | 19.15 (8 - 48) | 0.4894 |

CRP, C-reactive protein; PCT, procalcitonin; LDH, Lactate dehydrogenase; ALT, Alanine aminotransferase; PK, Prothrombin complex.

**Supplementary Table 7.** Analysis of the observed distribution of PhenoGraph clusters across clinical parameter-defined groups.

| Cluster | Disease severity<br>P values | Viremia<br>P values | SARS-CoV2-IgG status<br>P values | Oxygen need<br>P values | Steroid<br>treatment<br>P values | Chronic<br>disease<br>P values | Outcome<br>P values |
| --- | --- | --- | --- | --- | --- | --- | --- |
| C1 | 9.82469E-10 | 6.70053E-09 | 7.25622E-09 | 5.53078E-09 | 9.01886E-10 | 7.6966E-09 | 6.55857E-09 |
| C2 | 6.24061E-19 | 6.16635E-27 | 6.7457E-102 | 2.60388E-28 | 4.2181E-26 | 1.16983E-21 | 1.6289E-129 |
| C3 | 0.056787117 | 0.031879758 | 0.025540559 | 0.103800081 | 0.039535451 | 0.054414821 | 0.032339133 |
| C4 | 4.94484E-21 | 4.70244E-21 | 3.65038E-20 | 3.33757E-20 | 4.39116E-21 | 3.93243E-21 | 2.27658E-21 |
| C5 | 0.037769269 | 0.205805457 | 0.013978172 | 0.014115671 | 0.287612181 | 0.185839036 | 0.298050258 |
| C6 | 0.107393726 | 0.169741281 | 0.025309908 | 0.297765292 | 0.089839584 | 0.171999248 | 0.164256687 |
| C7 | 0.852296094 | 0.432611154 | 0.027118573 | 0.291099167 | 0.924229949 | 0.61966433 | 0.935662915 |
| C8 | 0.010428494 | 0.007242446 | 0.006279099 | 0.024407348 | 0.010367282 | 0.009634834 | 0.010389013 |
| C9 | 0.964356127 | 0.966709777 | 0.050672506 | 0.997375437 | 0.299360634 | 0.942940919 | 0.811646732 |
| C10 | 4.76455E-24 | 4.73336E-24 | 3.85027E-23 | 3.96539E-23 | 4.71891E-24 | 4.79459E-24 | 4.79458E-24 |
| C11 | 0.033692958 | 0.173035921 | 0.057367791 | 0.11260056 | 0.828275691 | 0.494646404 | 0.451895019 |
| C12 | 7.21691E-05 | 2.07415E-07 | 2.83626E-07 | 6.34108E-22 | 1.56775E-08 | 3.07716E-08 | 0.020981315 |
| C13 | 1.33951E-18 | 1.1605E-18 | 6.48612E-18 | 8.6982E-18 | 1.23499E-18 | 1.05436E-18 | 1.57293E-18 |
| C14 | 3.96969E-15 | 0.000866453 | 1.88733E-09 | 6.09675E-15 | 2.48072E-05 | 1.89865E-06 | 8.24868E-06 |
| C15 | 0.0921441 | 0.144607536 | 0.025917234 | 0.258672244 | 0.126054935 | 0.111460307 | 0.048732961 |
| C16 | 2.69559E-38 | 6.55861E-46 | 1.65959E-42 | 3.62433E-53 | 7.54469E-40 | 4.08116E-44 | 7.0591E-178 |
| C17 | 3.58485E-08 | 1.44336E-05 | 4.3292E-16 | 1.026E-09 | 4.69916E-06 | 4.41599E-07 | 5.45541E-10 |
| C18 | 0.01638769 | 0.001708367 | 0.039139449 | 0.044363228 | 0.015142306 | 0.000447192 | 0.019549737 |
| C19 | 0.034894632 | 0.537756332 | 0.09374095 | 0.205936535 | 0.197072037 | 0.812005299 | 0.196680606 |
| C20 | 2.25882E-11 | 1.83782E-10 | 1.16552E-27 | 3.71466E-18 | 2.88532E-10 | 3.54009E-27 | 2.97035E-30 |
| C21 | 1.0836E-68 | 1.0292E-52 | 6.6425E-106 | 4.6953E-134 | 2.09295E-52 | 1.00456E-42 | 4.71104E-24 |
| C22 | 3.15515E-09 | 5.01015E-09 | 1.87519E-06 | 2.1113E-07 | 3.59671E-11 | 2.18899E-08 | 4.57661E-07 |
| C23 | 0.447500686 | 0.670789196 | 0.023881369 | 0.727372625 | 0.234126333 | 0.544765353 | 0.217805037 |
| C24 | 3.7098E-65 | 4.47221E-46 | 1.34141E-42 | 2.98251E-79 | 2.91841E-42 | 3.82455E-39 | 9.85524E-21 |
| C25 | 0.325611881 | 0.808087094 | 0.45647612 | 0.599218214 | 0.216268288 | 0.622930226 | 0.160136923 |
| C26 | 6.33917E-17 | 4.22405E-15 | 1.93163E-32 | 1.02215E-16 | 4.78234E-22 | 4.26104E-15 | 4.54571E-15 |
| C27 | 3.3765E-08 | 1.92463E-08 | 1.94879E-11 | 1.32037E-07 | 3.42407E-08 | 2.61267E-09 | 2.13426E-08 |
| C28 | 0.723711804 | 0.548938267 | 0.081762179 | 0.854459924 | 0.461466554 | 0.655519801 | 0.625084789 |
| C29 | 4.53505E-09 | 9.69918E-10 | 2.17701E-11 | 1.8556E-08 | 4.39298E-09 | 7.73509E-10 | 2.4393E-09 |
| C30 | 1.58203E-31 | 7.78876E-25 | 1.94285E-35 | 2.58058E-27 | 2.63203E-41 | 7.73609E-20 | 6.85559E-77 |
| C31 | 4.86534E-31 | 2.00966E-44 | 1.179E-135 | 1.1186E-108 | 1.71782E-41 | 3.56777E-37 | 6.63092E-24 |
| C32 | 1.95728E-09 | 1.57979E-09 | 7.93393E-10 | 8.35178E-09 | 1.95019E-09 | 6.27335E-12 | 1.99177E-09 |
| C33 | 2.9461E-17 | 2.95618E-17 | 1.65472E-16 | 1.76962E-16 | 2.71956E-17 | 2.79302E-17 | 2.9599E-17 |
| C34 | 0.233957169 | 0.04317915 | 0.026294868 | 0.005533287 | 0.027787029 | 0.000759428 | 0.180994372 |
| C35 | 0.002731175 | 0.002906441 | 0.002823929 | 0.007648985 | 0.002732727 | 0.001336849 | 0.002789391 |
| C36 | 0.105886144 | 0.081275019 | 0.032405698 | 0.171898519 | 0.076151088 | 0.005478345 | 0.034644632 |

